# The impact of mobility network properties on predicted epidemic dynamics in Dhaka and Bangkok

**DOI:** 10.1101/2021.02.07.21250586

**Authors:** Tyler S. Brown, Kenth Engø-Monsen, Mathew V. Kiang, Ayesha S. Mahmud, Richard J. Maude, Caroline O. Buckee

**Affiliations:** Massachusetts General Hospital, Infectious Diseases Division; Harvard T.H. Chan School of Public Health; Telenor Research; Stanford University School of Medicine, Department of Epidemiology and Population Health; University of California, Berkeley, Demography Department; Mahidol-Oxford Tropical Medicine Research Unit, Faculty of Tropical Medicine, Mahidol University; Centre for Tropical Medicine and Global Health, Nuffield Department of Medicine, University of Oxford

**Keywords:** Cities, human mobility, mobile phone data, SARS-CoV2 dynamics

## Abstract

Properties of city-level commuting networks are expected to influence epidemic potential of cities and modify the speed and spatial trajectory of epidemics when they occur. In this study, we use aggregated mobile phone user data to reconstruct commuter mobility networks for Bangkok (Thailand) and Dhaka (Bangladesh), two megacities in Asia with populations of 16 and 21 million people, respectively. We model the dynamics of directly-transmitted infections (such as SARS-CoV2) propagating on these commuting networks, and find that differences in network structure between the two cities drive divergent predicted epidemic trajectories: the commuting network in Bangkok is composed of geographically-contiguous modular communities and epidemic dispersal is correlated with geographic distance between locations, whereas the network in Dhaka has less distinct geographic structure and epidemic dispersal is less constrained by geographic distance. We also find that the predicted dynamics of epidemics vary depending on the local topology of the network around the origin of the outbreak. Measuring commuter mobility, and understanding how commuting networks shape epidemic dynamics at the city level, can support surveillance and preparedness efforts in large cities at risk for emerging or imported epidemics.

## 2 Introduction

Densely populated cities are uniquely vulnerable to infectious disease epidemics [1]. The distribution and connectivity of human populations within cities plays a key role in spreading outbreaks when they occur, but the factors determining how the vulnerabilities translate into epidemic dynamics are still unclear, and this has important implications for surveillance and preparedness. Local SARS-CoV-2 epidemics have thus far exhibited substantial diversity in their scale and trajectory between cities [2], for example, and the factors behind differential city-level risk of epidemic propagation (for both SARS-CoV-2 and other emerging or endemic pathogens) are still incompletely understood [3, 4]. Likewise, our ability to predict the spatial distribution of disease activity within cities during epidemics (for example, by neighborhood or hospital catchment area) is constrained by lack of informative data, including geolocated or spatially-resolved epidemiological data and reliable data on human movement in cities. Improved strategies are needed for *a priori* stratification of epidemic risk in large cities, including strategies that can estimate epidemic potential at the city level and identify local determinants of epidemic risk within cities.

Commuter mobility is an important factor in the local dispersal of directly transmissible pathogens (for example, influenza [5] and for SARS-CoV-2 [6] and differences in commuter mobility patterns predict divergent epidemic dynamics between cities [7]. Mobile phone call detail records (CDRs) have become an important tool for estimating human mobility, and the utility of CDR data for modeling infectious disease dynamics has been demonstrated in multiple contexts [8, 9] We propose that CDR-derived approximations of commuter mobility networks in cities can inform city-level predictions of epidemic risk, and we hypothesize that both node-level and higher-order properties of these networks influence the temporal and spatial trajectory of city-wide epidemics.

In this study, we employed aggregated CDR data to estimate commuter mobility networks in Bangkok, Thailand and Dhaka, Bangladesh and applied a stochastic model for propagation of a directly transmitted infection (similar to SARS-CoV-2 or influenza) on these networks. These two cities capture some of the substantial diversity in population density, spatial organization, transportation networks, and socioeconomic conditions observed across “megacities” (typically defined as cities and associated urban agglomerations with *>* 10 million total residents), and are thus well-suited for studying differences in epidemic dynamics between large cities. We find important differences between commuter mobility networks and predicted spatiotemporal trajectories of epidemics in each city. We report metrics on spatial distribution of population density in megacities, including Dhaka and Bangkok, to contextualize these findings. Our results support the use of mobile phone user data for evaluating city-level susceptibility to emerging or imported epidemics and for identifying locations within cities where local topology of the commuter network is expected to facilitate epidemic propagation.

## 3 Methods

### 3.1 Spatial heterogeneity in population density across megacities

To contextualize our analysis of city-level mobility and epidemic dynamics in Dhaka and Bangkok, we calculate multiple metrics summarizing the spatial distribution of population density across Dhaka and Bangkok [10], using data from the WorldPop project [11]. These include both spatially-naïve metrics (Gini coefficient, entropy, and relative standard deviation) and the “spatial dispersion index”, a spatially-explicit metric characterizing heterogeneity in population density across a given area (the “spreading index” in [10]). Additional information on these metrics is provided in the Supplemental Information. To control for the different sizes and shapes of the study areas in each city, we calculate each metric over concentric circles centered on the most densely populated grid square in each city, and report the metrics by the radius *r* of each circle. For context, we report these metrics for 19 additional megacities located in low- and middle-income countries (Bogota, Columbia; Cairo, Egypt; Guadalajara and Mexico City, Mexico; Ho Chi Minh City, Vietnam; Hyderabad, Jaipur, Kolkata, Mumbai, and Pune, India; Istanbul, Turkey; Johannesburg, South Africa; Karachi and Lahore, Pakistan; Brazzaville-Kinshasa, Congo-Democratic Republic of Congo; Lagos, Nigeria; Manila, Philippines; Sao Paulo, Brazil; and Tehran, Iran).

### 3.2 Commuting networks estimated from mobile phone user data

Daily commuter flux between locations in Bangkok and Dhaka was estimated using aggregated, anonymized CDR data for 4.3 and 18.9 million average daily mobile phone subscribers in the Bangkok Metropolitan Region (BMR) and Dhaka Statistical Metropolitan Area (DSMA), respectively (Supplemental Figure S1). The BMR is an administratively-defined region that includes Bangkok and five surrounding provinces, covering 7762 *km*^2^ with an estimated population of approximately 15.9 million (2048 *persons/km*^2^). The DSMA is composed of Dhaka and several surrounding administrative units covering 1353 *km*^2^ with an estimated population of approximately 21 million (15521 *persons/km*^2^). We collected CDR data over 81 consecutive days (1 August to 19 October, 2017 in Bangkok, and 1 April to 21 June 2017 in Dhaka), restricted to the mobile network towers within each administrative region (BMR or DSMA), and excluded major national and religious holidays. We aggregated data over 500 m x 500 m grid squares and used squares with at least one mobile phone tower in service during the data collection period to define the set of nodes ℒ = {*L*_1_, *L*_2_, …, *L*_*i*_} in the commuting network. We used Voronoi polygons to define the catchment area around each node and used estimated population maps from WorldPop [11] to assign the population in each catchment area *N*_*i*_ for each node *L*_*i*_ ∈ℒ.

To estimate daily trip counts between nodes, we identify the mobile network tower used for the majority of each user’s calls (the “most-visited” node) for each of two consecutive 24-hour periods. We assume that the most-visited node during the first 24-hour period and the most visited node during the following 24-hour period represent the origin and destination, respectively, of a single daily trip between the areas serviced by the tower at each node. In brief, the mean number of raw, unweighted trips originating from *L*_*j*_ and terminating at *L*_*k*_, averaged over each daily observation in the data collection period, is denoted 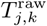. We estimate the population-weighted number of trips originating from node *L*_*j*_ and terminating at node *L*_*k*_ by apportioning the total population in *N*_*j*_ by the relative proportion of all raw trips counts that originate at *L*_*j*_ and terminate at node *L*_*k*_.

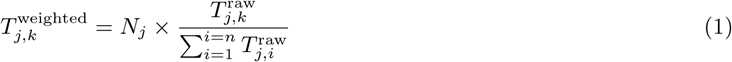

where *n* is the total number of nodes in ℒ. 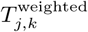 thus represents the proportion of the population at node *L*_*j*_ that regularly travels between *L*_*j*_ and *L*_*k*_. We used the resulting trip counts to construct an *n* × *n* origin-destination matrix ℳ_OD_ where each entry 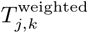 equals the weighted mean number of daily trips between nodes *L*_*j*_ and *L*_*k*_ during the data collection period (and rows and columns are indexed by *j* and *k*, respectively).

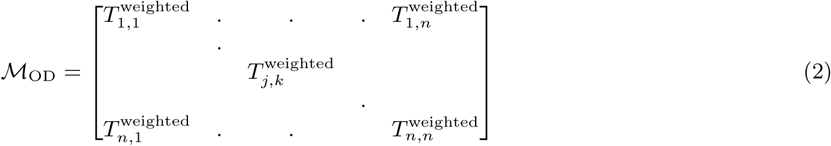

We consider two separate versions of ℳ_OD_: 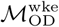, which includes only trips originating on the last day of the weekend and terminating on the first day of the conventionally observed work week (Monday in Bangkok and Sunday in Dhaka); and 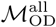, which includes trips originating on all days in the data collection period. Analysis using 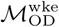 is motivated by the assumption that the most visited location on weekend days is likely to represent a user’s home location, and the most visited location during the first day of the work week is likely to be a user’s work location, such that 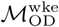 is expected to capture commuter mobility between home and work locations. 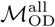 captures movement between frequently-visited locations and, although there is no expectation that these are either home or work locations, provides a proxy measurement for aggregate daily human movements between locations within each city. Entries 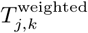 in 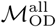 are calculated using all 81 available origin days in the data collection period and 8 observed weekend-weekday transitions in the data collection period for 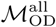. 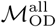 and 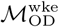 are highly similar to one another (Mantel test *p <* 0.005) and the mobility networks estimated from the two matrices share are very similar with respect to network community structure and centrality measures (Supplemental Figures S2-S5), i.e. we observe the same overall mobility patterns between the two methods of estimating mobility flux. We use 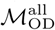 for the primary analyses that follow, given the large number of days in this data collection period, which is expected to provide more informative data compared to 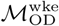, which is derived from a much smaller number of observations. Additional data processing procedures and analyses using 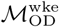 are reported in the Supplemental Information.

### 3.3 Characterization of commuter mobility networks

Using the edge-weighted, directed network specified by the final origin-destination matrices (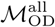 and 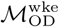), we calculate multiple node-level metrics describing local network topology, including degree (total number of incoming and outgoing edges for each node) and strength (the sum of all incoming or outgoing edge weights given by 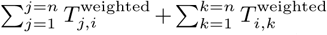 for node *L*_*i*_). We also calculate eigenvector centrality, defined as the largest positive eigenvector for the network adjacency matrix, which prior work has identified as an important indicator for spreading power in networked epidemics [12]. Following Brockmann *et al*.[13], we calculate the effective distances between node, based on estimated connectivity rather than geographic distances, as 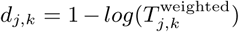 over the shortest estimated path between nodes *L*_*j*_ and *L*_*k*_. We inferred the community structure of the commuting network, i.e., the size and membership of distinct modular subnetworks within the larger network, using InfoMap [14, 15, 16]. Briefly, InfoMap estimates community structure by minimizing the length of the Huffman code [17] descriptor for the path of a random walk across a given network [14].

### 3.4 Stochastic modeling of epidemic propagation on commuter mobility networks

We use a stochastic metapopulation model to estimate the propagation of a directly-transmissible immunizing infection with susceptible-exposed-infected-recovered (*SEIR*) dynamics over the city-level commuting networks in Dhaka and Bangkok. Although the duration of protective immunity acquired during SARS-CoV2 infection is still uncertain [18], we assume that recovered individuals are not susceptible to infection, at least for the relatively short periods of time considered in our simulations. The framework of the model follows [19] and [20], with the initial population of susceptible individuals distributed into origin-destination compartments specified by 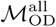 and 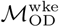. We assume that all individuals are susceptible to infection at time *t* = 0.

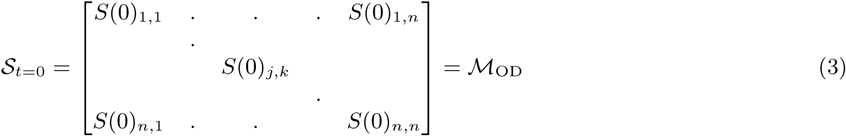

The number of exposed, infected, and recovered individuals in each corresponding compartment at time *t* are given by the *n* × *n* matrices *ℰ*, ℐ, and ℛ, with entries *E*_*j,k*_,*I*_*j,k*_, and *R*_*j,k*_, respectively. Time-steps are days, and new exposures (*S* → *E* transitions) result from interaction with infected individuals at either the origin or destination, with force of infection *λ*^origin^ and *λ*^destination^, respectively.

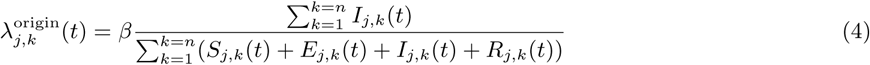

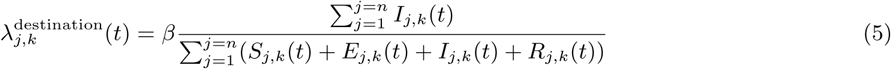

where *β* is a constant specifying the risk of infection (converted from the daily rate of infection estimated in [20]). 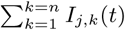 and 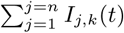 are the number of infected individuals to which the *S*_*j,k*_(*t*) susceptible individuals are exposed to in the origin and destination nodes *j* and *k*, respectively. For each time-step, the number of susceptible individuals *S*_*j,k*_(*t*) who are exposed is the sum of two draws from two different binomial distributions with probabilities *λ*^origin^ and *λ*^destination^. Infected individuals are parsed into reported and unreported cases, with *u* denoting the relative infectiousness of reported versus unreported infections. The model does not consider births or deaths, such that the total populations across compartments are invariant over time and, for all times *t*, the denominators in Equations (4) and (5) equal 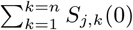 and 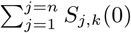, respectively. Progression to infectiousness (*E* → *I* transitions) and recoveries (*I* → *R* transitions) occur with fixed probabilities *η* and *γ*.

### 3.5 Modeling parameter selection and sampling

Base parameters for the stochastic SEIR model were chosen from parameters inferred for a similar model using COVID-19 disease report data from China [20]. We sample parameters from across the 95% credible interval for each parameter *β, η, γ*, and *u* as estimated by Li et al [20] (specifically, estimates derived from data collected during early, pre-intervention stages of the epidemic) using Latin hypercube sampling. For some analyses *β, η, γ*, and *u* are set to fixed values of interest.

## 4 Results

### 4.1 City structure differences between Bangkok and Dhaka

Calculated values for the Gini coefficient and relative standard deviation, which both increase with increasing statistical dispersion, are higher across Dhaka versus Bangkok (Figure 1), consistent with greater heterogeneity in population density in Dhaka. Likewise, calculated entropy values, which decrease with increasing statistical dispersion, are lower across all spatial scales (i.e. concentric circles of radius *r*) in Dhaka. The spatial dispersion index is lower in Dhaka for all values of *r*, indicating that cells with higher population density are distributed over smaller areas in Dhaka versus Bangkok. The spatial distributions of population density in Dhaka and Bangkok span the range of observations for other large cities in low- and middle-income countries (Figure 1, grey lines). The spatial distribution of population density in Dhaka is generally more heterogeneous and concentrated over smaller spacial scales versus other cities, whereas the spatial distribution of population density in Bangkok is “flatter” and distributed over larger areas versus other cities (Figure 2A).

**Figure 1:**
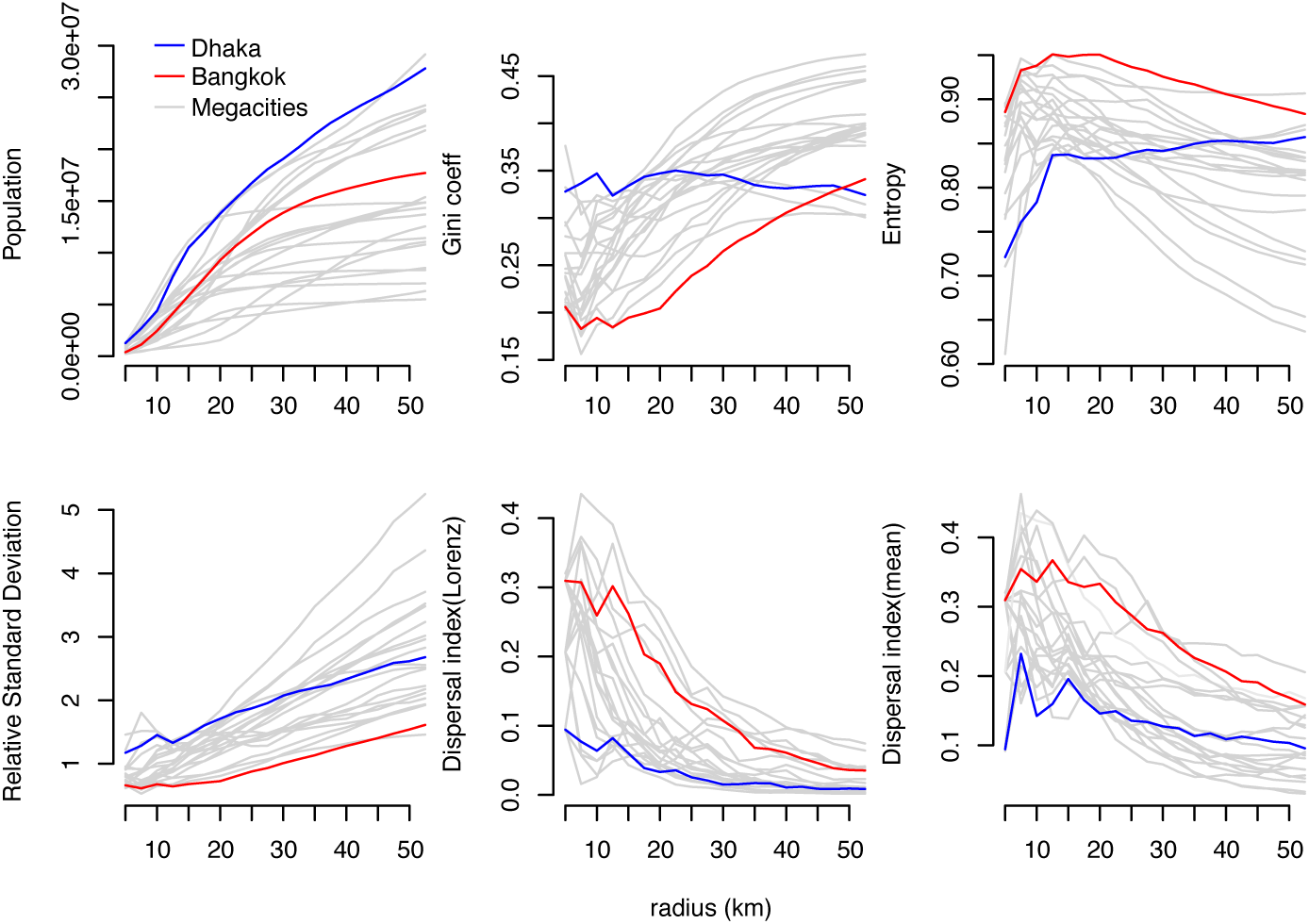
Spatial distribution of population density in Bangkok and Dhaka. Total population, three spatially-naïve metrics of population density (Gini coefficient, entropy, and relative standard deviation), and one spatially-explicit metric of population density (spatial dispersion index) are shown for Bangkok (red lines), Dhaka (blue lines), and 19 additional megacities (grey lines). Each metric is estimated over concentric circles centered on the WorldPop cell with highest population density in each city (see Figure 2). The dispersal index is estimated using two different threshold values, the mean of all population density values and the “Loubar” threshold value derived from the Lorenz curve of population density values, as described in [10].

**Figure 2:**
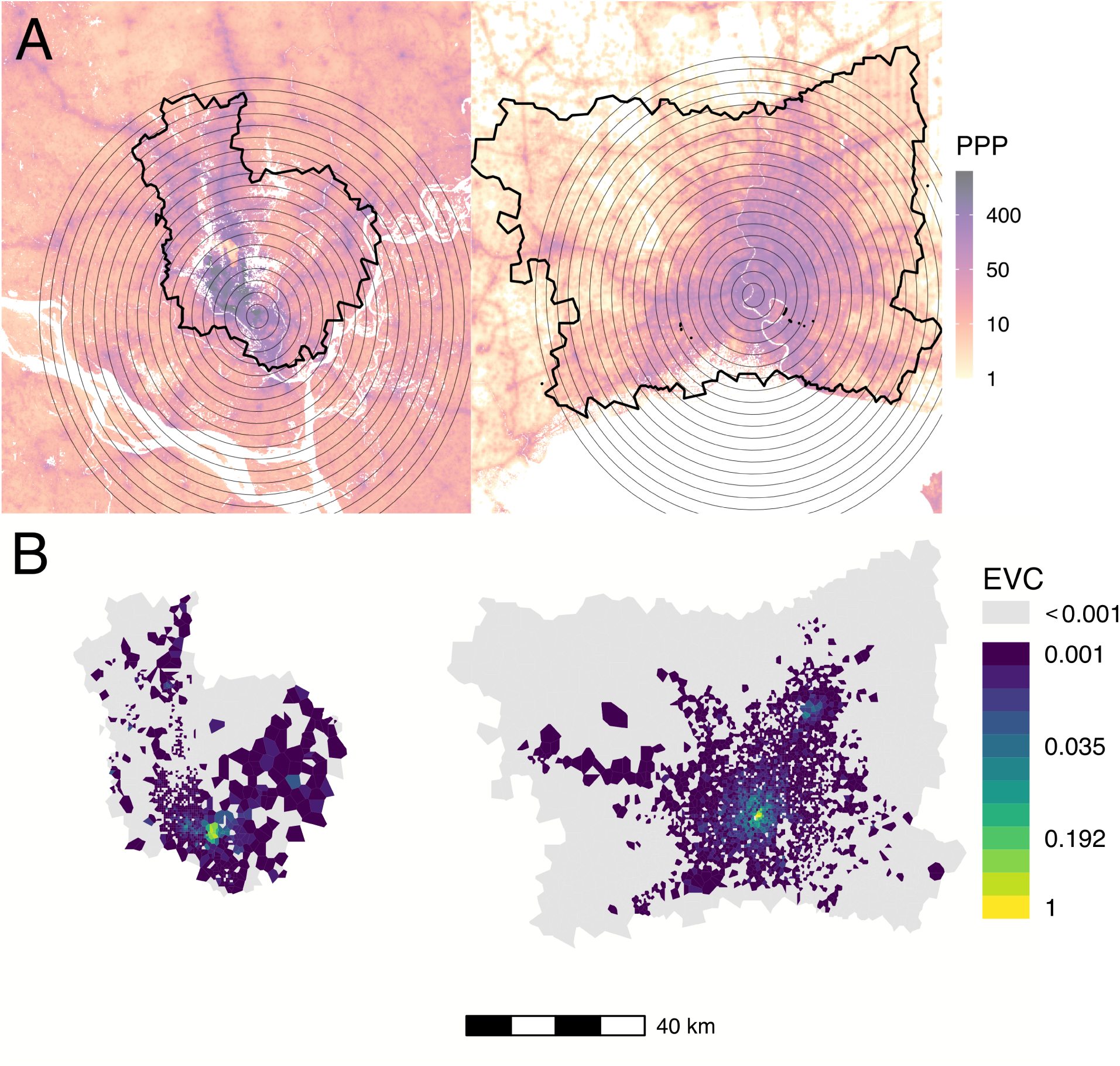
Population density, mobile data catchment areas, and eigenvector centrality in Dhaka (left) and Bangkok (right). (A) Population density in persons per pixel (PPP) from WorldPop database, overlaid with concentric circles used for spatially-restricted measurements of population density in Figure 1 (grey circles) and the outline of the mobile service tower-associated catchment areas used to estimate commuter mobility in each city. (B) Eigenvector centrality by node for Dhaka and Bangkok for the mobility network specified by 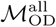.

### 4.2 Network properties and community structure in Dhaka and Bangkok

Node-level properties demonstrate important differences in overall connectivity and network density between Dhaka and Bangkok. The node degree distribution in Dhaka is concentrated around higher values, with most nodes sharing edges with all or almost all other nodes, with a corresponding network edge density (the ratio of the number of edges and the number of possible edges in a network) of 0.82; the degree distribution in Bangkok is less concentrated at higher values, with a corresponding network edge density of 0.11 (Supplemental Figure S6). The distribution of node strengths is more widely dispersed and has higher maximum values in Dhaka compared to Bangkok.

Network centrality in both cities is concentrated within more highly populated areas. Eigenvector centrality values are highest in centrally-located areas with high population density in both Bangkok and Dhaka; in Bangkok, we observe a second focus of nodes with high eigenvector centrality values near a known transportation hub in the northeast area of the BMR (Figure 2B).

Network community structures are distinctly different in Dhaka and Bangkok. Figure 3 maps the ten largest network communities in either Dhaka or Bangkok, inferred from the commuter network specified in 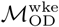. In this context, network communities represent subnetworks of nodes in the commuting network that are more strongly connected to each other compared to other nodes. Network communities in Bangkok (Figure 3B) are geographically contiguous and constrained by geographic barriers (for example, the Chao Praya River); with some exceptions, communities in Dhaka are mostly discontiguous and relatively unconstrained by geographic barriers (Figure 3A) or distance (Figure 3C). The geographic contiguity of network communities in Bangkok is observed even after down-sampling to a smaller set of nodes within a restricted geographic area (Figure S7), indicating that differences in community structure observed between Dhaka and Bangkok are likely not related to differences in connectivity to unobserved nodes not included in the data catchment area.

**Figure 3:**
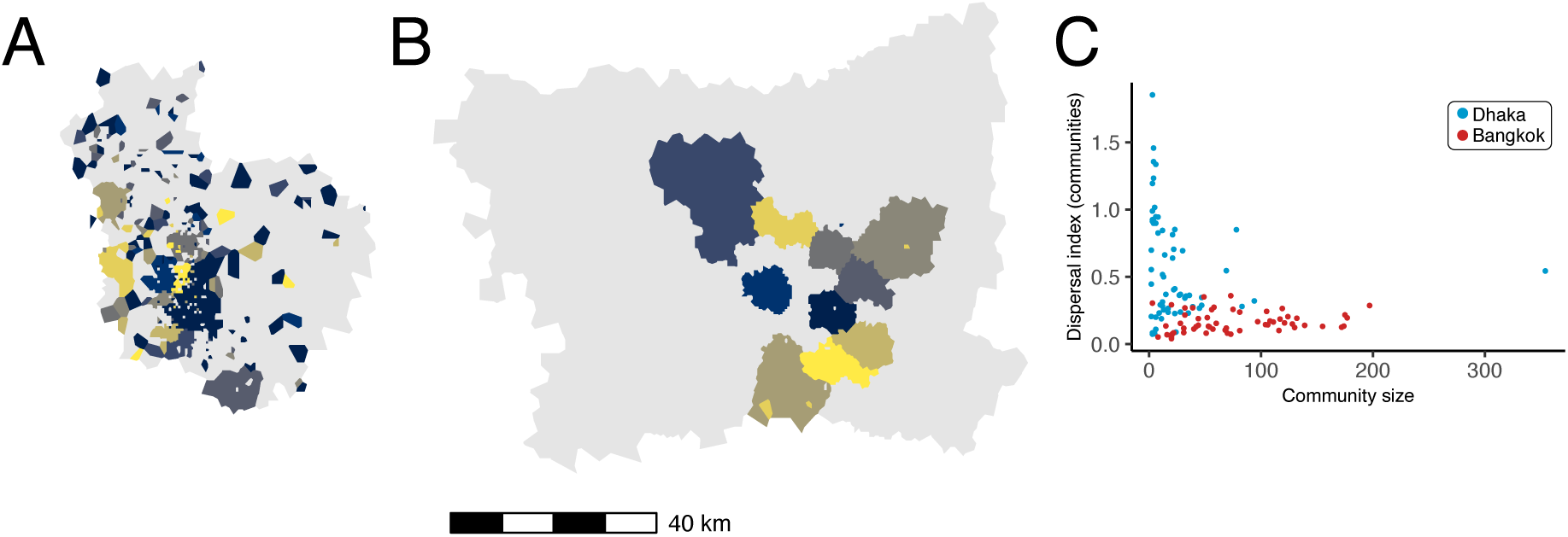
Commuter mobility network structure in Dhaka and Bangladesh. The ten communities with largest membership (by number of nodes) are mapped for (A) Dhaka and (B) Bangkok. Panel (C) plots community membership size (by number of nodes) versus the spatial dispersion index calculated for the nodes in each community (blue: Dhaka, red: Bangkok). Colors for polygons in (A) and (B) are arbitrary.

### 4.3 Propagation of simulated epidemics: synchrony and predictability

In simulated epidemics of a directly-transmitted pathogen with SEIR dynamics, initialized using parameters inferred for SARS-CV2 transmission, we observe highly synchronized epidemic dynamics in Dhaka (Figures 4A & 4B), with all nodes across the city reaching their highest number of predicted infections within a short, synchronized period of time. Epidemic propagation is less synchronized in Bangkok and nodes located in the peripheral areas of the BMR, specifically in the west and northeast, exhibit delayed epidemic peaks compared to nodes in central Bangkok or the western BMR (Figures 4C & 4D). These results are consistent across a wide range of modeling parameters and are observed regardless of how the origin node for each simulation is chosen (either from a uniform distribution or population-weighted multinomial distribution, Figure S8).

**Figure 4:**
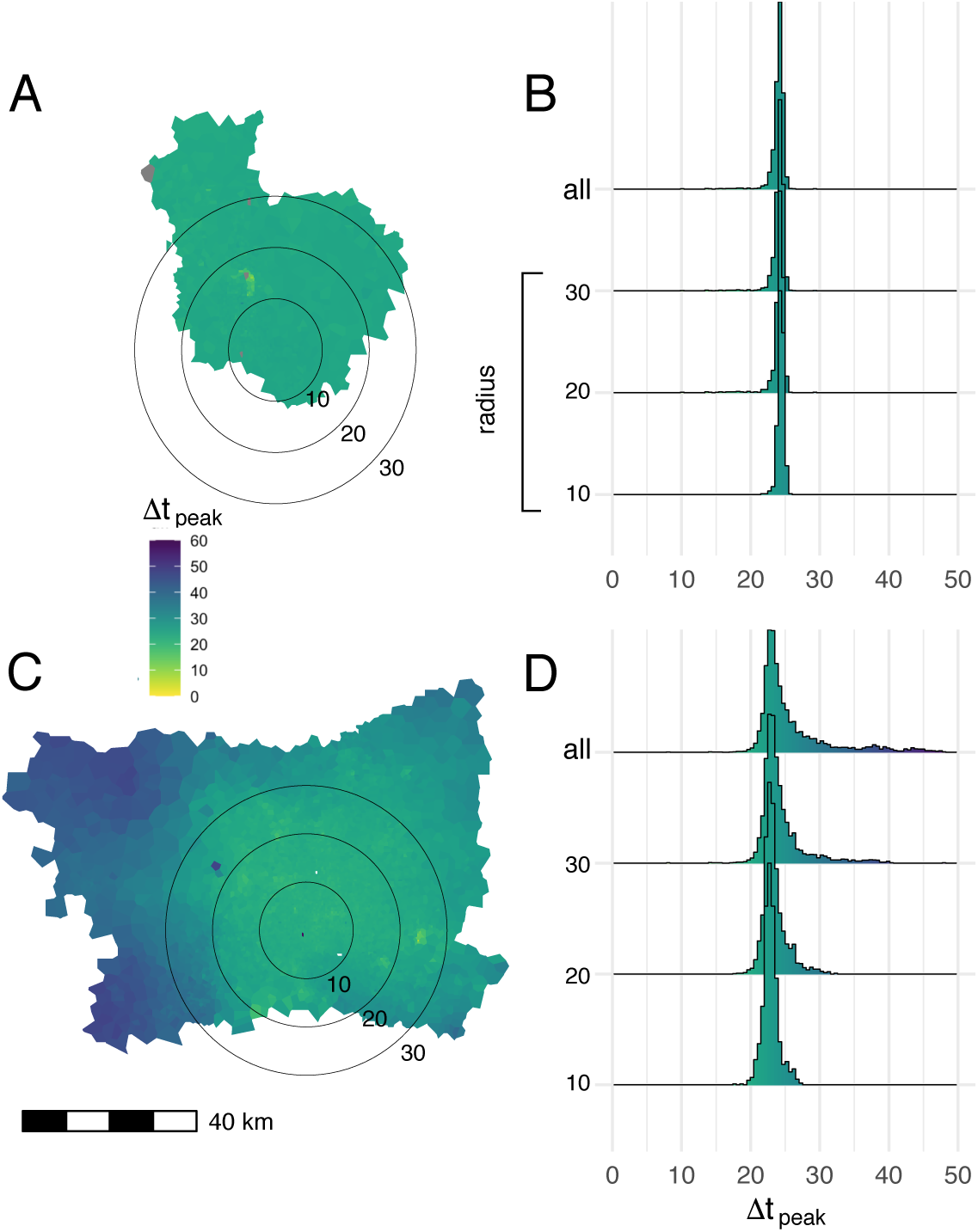
Mean time to epidemic peak for each node in the commuting network. Map polygons are colored according to the time lag in days between the time to epidemic peak in a given node and the earliest peaking node on the map, Δ*t*_peak_ (A:Dhaka, C: Bangkok). Results from 1,000 independent SEIR simulations, using parameters drawn via Latin hypercube sampling (as described in *Methods*), are shown. The seed node for each simulation is drawn from a multinomial distribution, where the probability that a given node is chosen as the seed is proportional to the total population in the catchment area (Voronoi polygon) around that node. Histograms show the distribution of epidemic peak times over different geographic areas in Dhaka (B) and Bangkok (D), including nodes within concentric circles centered on most densely populated node in each city (radii: 10,20, and 30 km) and the entirety of both study areas (“all” in panels B and D).

Simulated epidemics initialized on the Bangkok commuting network exhibit distinct wave-like dispersal away from an epidemic origin (Figure 5B). Early in the simulated epidemics, the number of infected individuals in a given node is negatively correlated with geographic distance from the origin node. This correlation becomes less distinct and later reverses and becomes positive as the wave of epidemic dispersal moves outward from the epidemic origin. Simulated epidemics on the Dhaka commuter network do not exhibit any wave-like features and, unlike Bangkok, propagation of the epidemic is largely unconstrained by distance (Figure 5A) at all time points. These features are observed regardless of how origin nodes are selected (either from a population-weighted multinomial distribution or a uniform distribution, Figure S9), and are also observed if the mobility network is specified by 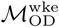 rather than 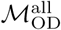 (Figure S10). Similar findings are observed when the mobility network is restricted to nodes within a specified distance from the epidemic origin (60 km in Figure S11 and 30 km in Figure S12).

**Figure 5:**
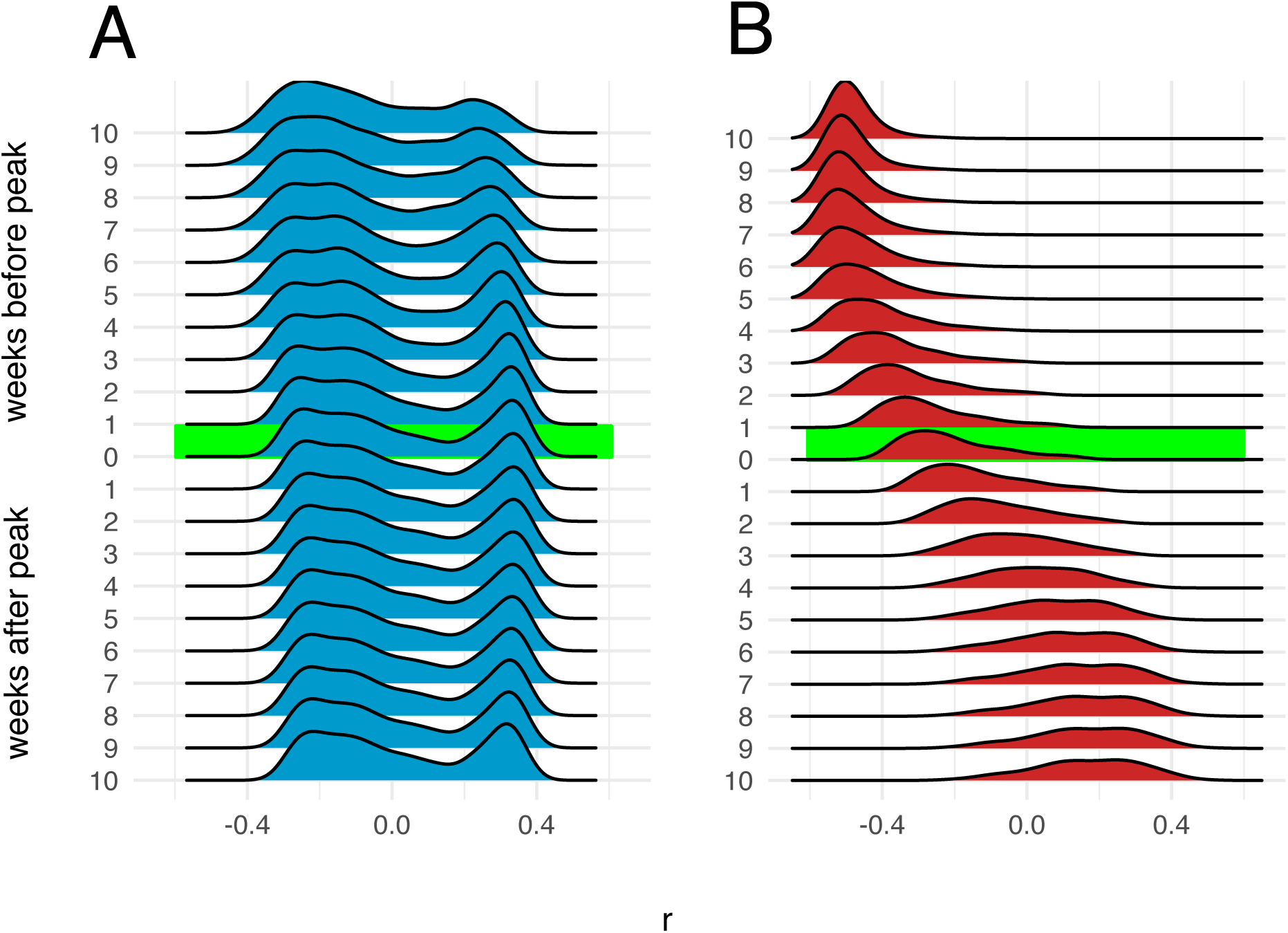
Correlation between number of infections and distance from epidemic origin node over time. Distributions of Pearson’s coefficients for correlation between number of infections in each node and nodes’ respective distances in km from the origin node, obtained from 1,000 independent SEIR simulations, are shown at different time points during simulated epidemics. The origin node for each simulation is chosen from a multinomial distribution weighted by the total population in the catchment area (Voronoi polygon) around each node. A: Dhaka, B: Bangkok.

### 4.4 Local network topology influences epidemic trajectory

We next examined dynamics of simulated epidemics originating from nodes with high and low local connectivity, as measured by their eigenvector centrality values. In simulated epidemics propagating on the Bangkok commuter network, epidemic speed (measured as the time to peak number of infections in the entire network) differs depending on the eigenvector centrality of the origin node (Figure 6): specifically, simulated epidemics originating at nodes with lower eigenvector centrality values are slower, with later epidemic peak times, compared to those originating at nodes with high eigenvector centrality (mean time to epidemic peak, 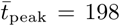 and 222 days for nodes with eigenvector centrality values in the tenth and first deciles, respectively). For simulated epidemics initialized on the Dhaka commuter network, we observe a smaller difference in time to epidemic peak between simulations seeded at nodes with low versus high eigenvector centrality values (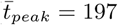 and 203 days). Maximum epidemic size, i.e. the estimated number of infected individuals at the epidemic peak, also varies by eigenvector centrality of the origin node with smaller differences observed in Dhaka compared to Bangkok (Figure S13). The degree, strength, and eigenvector centrality of the origin node are more strongly correlated with mean epidemic arrival time (time to first infection in a given node, averaged over all nodes in the network) in Bangkok compared to Dhaka (Supplemental Figure S6). Geographic distance and effective network distance (a measure of mobility-based connectivity between nodes) from the origin node are positively correlated with epidemic arrival time in Bangkok (Supplemental Figure S14); in Dhaka, epidemic arrival time is positively correlated with effective network distance, but is less consistently correlated with geographic distance. In both Dhaka and Bangkok, for epidemics simulated with a wide range of model parameters and seeded at random nodes across each network, epidemic arrival time is negatively correlated with eigenvector centrality of non-origin nodes (Supplemental Figure S14).

**Figure 6:**
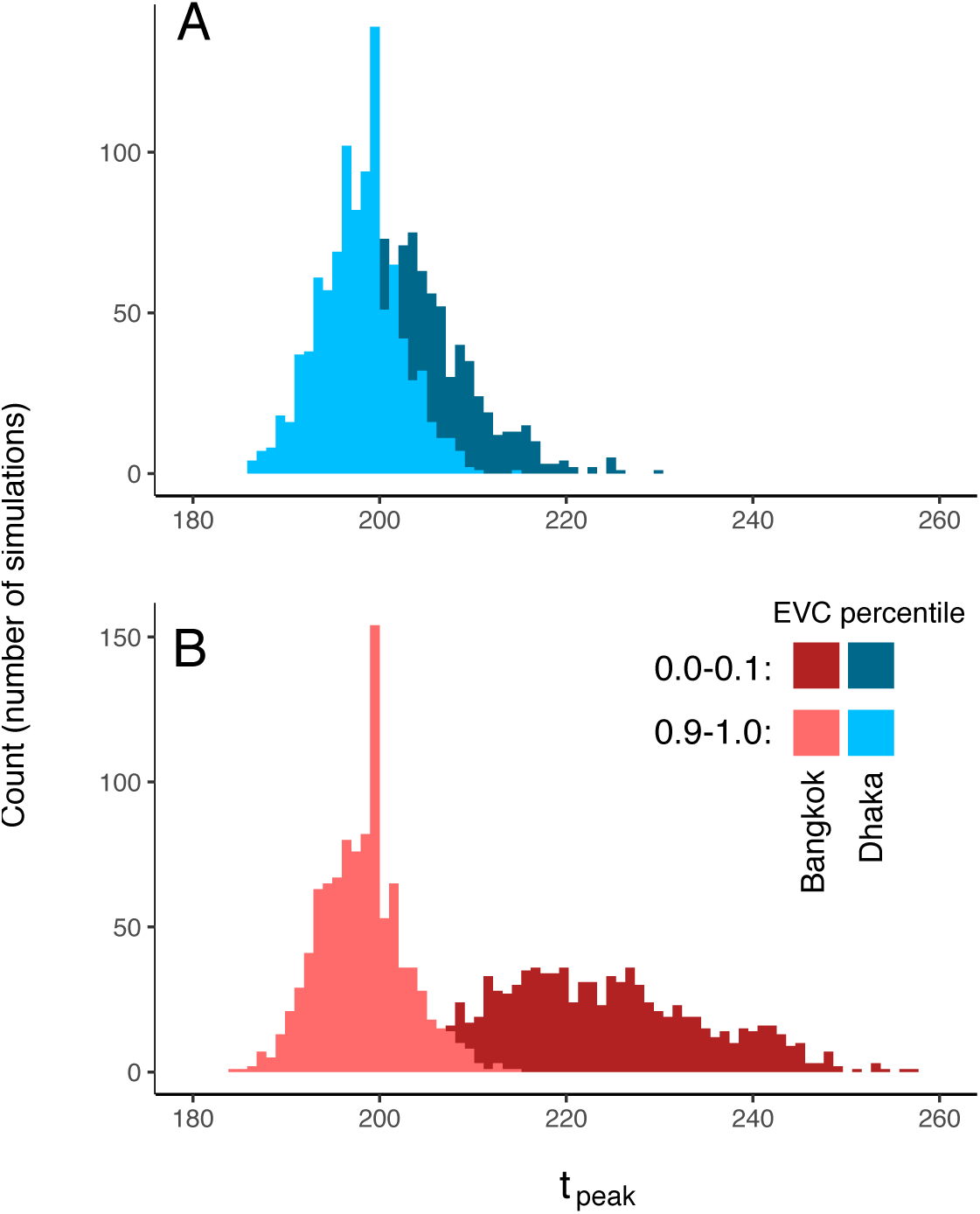
Epidemic dynamics and eigenvector centrality at the origin node. Distributions for time (in days) to epidemic peak for are shown for simulated epidemics seeded at nodes with high and low eigenvector centrality values (in the first and tenth deciles, respectively). A: Dhaka; B: Bangkok. Distributions are for 1,000 simulated epidemics with *β* = 1.12*days*^−1^.

## 5 Discussion

Epidemic dynamics in cities will depend on a range of factors including the connectedness and distribution of its populations. The timing, appropriateness, and efficacy of disease control interventions undertaken in response to ongoing or potential epidemics are clearly central determinants in how epidemics unfold in different cities, and may supersede the impact of many other city-level properties that might modify epidemic risk. Nevertheless, understanding how observable properties of cities shape their intrinsic vulnerability to epidemic propagation remains an important scientific and public health priority. Here, we focus on mobility, acknowledging that mobility itself is closely intertwined with other important factors such as population density, economic conditions, and public infrastructure for transportation, public health, and social support.

Megacities as a group exhibit wide variation across these potential determinants of epidemic dynamics. Building on prior, foundational work on city-level mobility and epidemic risk [7], we have focused our analysis on Bangkok and Dhaka, two cities with distinctly different economic and social conditions that also capture the wide variation in spatial population structure across megacities (Figure 1). The resulting analysis provides informative comparative findings, delineating key differences in underlying mobility networks and predicted epidemic dynamics in these two example cases. We also find important associations between characteristics of city-level mobility networks and multiple important epidemic features, including the synchrony, spatial dispersal, and size of simulated epidemics.

CDR-estimated mobility networks in Dhaka and Bangkok exhibit distinctly different network community structures. Network communities (i.e. modular subnetworks of nodes with relatively higher shared connectivity) in Bangkok are largely contiguous and geographically constrained, suggesting that mobility in the BMR is highly local and that daily movements of many individuals are limited to or organized around specific geographic areas (for example, neighborhoods or neighboring cities such as Nonthaburi). Communities in Dhaka are distinctly non-contiguous, suggesting movement within the DMSA is less local and not constrained by geography. Strong geographic contiguity within communities is observed in the BMR data even after restricting our analysis to a small area in central Bangkok, indicating that the geographically disorganized network communities observed in Dhaka are likely not an artifact of unmeasured connectivity with unobserved nodes outside the DMSA study area (Supplemental Information).

Simulated epidemics in Dhaka are tightly synchronized over time and space, unlike Bangkok, where simulated epidemics propagate as distinct spatial waves that arrive and peak later in outlying parts of the BMR, similar to prior observations on influenza epidemics in the United States [21]. These differences are robust over different spatial scales in each city (Figures 4B & 4D), indicating that the synchronized epidemics in Dhaka are not simply an artifact of the relatively smaller geographic size of the DMSA. This is an important alternative explanation to consider. Given a sufficiently large catchment area around the DMSA, we would expect to see delayed epidemic peaks at outlying or weakly connected locations; indeed, using our modeling approach, this would be an expected result for simulated epidemics propagated on any mobility network in which certain nodes have weaker connectivity to the network as whole (due to distance or other factors). Still, the observations in this study have potentially important implications for public health decision-making in the BMR and DMSA. Specifically, for Dhaka, these results may suggest that resources for testing and case detection (for example, PCR-based testing for SARS-CoV2) should be deployed as early and widely as possible during an epidemic, and that geographically-restricted testing (for example, at assumed transmission “hot spots”) could result in large numbers of infections going undetected.

Several other considerations are important for contextualizing our findings. Estimating mobility from mobile phone user data requires the use of multiple simplifying assumptions, some that are inherent to all estimates made from this kind of data, some that are essential to protect the privacy of mobile data users, and some that are specific to our study. Most important, we assume that two locations, (1) the node where a user places the majority of calls during one 24-hour period and (2) the node where the same user places the majority of calls during the subsequent 24-hour period, represent the origin and terminus of a single trip by one user. This assumption has potential shortcomings and the resulting origin-destination matrix may not fully capture daily movement. Numerous alternative approaches for estimating daily commuter movement from CDR data have been proposed and examined (for example, [22, 23, 24]) but as yet there is no consensus on which of these approaches provides the most appropriate informative data for modeling epidemic dynamics. How origin-destination matrices derived from CDR data compare with census-derived estimates [25], and how the use of these data sources influences models of epidemic dynamics [19, 26], are important research questions. We also note that the entries in 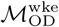 and 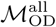 are scaled using estimated population data from the WorldPop project, and that these estimates have important sources of bias and variance as well [11]. We also acknowledge that mobility patterns in cities are dynamic and that analyses based on data aggregated over discrete time periods may not fully capture variability in mobility patterns over time. Likewise, population distributions and city structure may change rapidly in megacities and mobility data collected in these environments may become outdated relatively quickly. Lastly, by considering only trips between locations within the BMR and DSMA, we do not account for connectivity between these areas and more distant or outlying locations; connectivity with areas outside the BMR and DSMA is expected to not only drive epidemic dispersal into surrounding locations but also influence spatiotemporal epidemic dynamics within the central urban areas.

However, for multiple reasons, this method for estimating movement patterns is expected to capture important features of the mobility networks in each city, including features that are informative for modeling infectious disease dynamics. By using this approximation, and assuming the frequency of mobile phone use correlates to time spent in a given location, we capture the two most likely locations for each user, each day. Aggregated over millions of individual users, and averaged over an extended data collection period, we are able to obtain estimates for average connectivity between locations in a city based on large numbers of total observations. In addition, several intuitive findings from our analysis of the mobility networks specified by 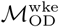 and 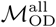, including higher eigenvector centrality values at geographic points with higher expected connectivity (for example, city centers and transportation hubs), support our approach to estimating city-level mobility patterns from aggregated CDR data.

Multiple limitations of our modeling approach are important to consider. The stochastic model used in this study assumes that mixing is homogeneous in each compartment and assumes that co-location in the same node is an adequate proxy for person-person interaction. Also, this model does not account for potential differences in the average time spent in each location (“dwell time”), which is likely to vary across locations resulting in differences in average daily force of infection by location, nor does it account for geographic heterogeneity in age or household structure. Additional studies, comparing directly observed person-person interactions (for example, via Bluetooth handshake [27] or RFID sensors [28]) to mobility trajectories estimated from CDR and other passively-collected mobile data sources, are needed to better understand the limits of CDR-based mobility estimates in this context. Lastly, we initialize our model with a homogeneously (and entirely) susceptible population, and as such this approach is poorly suited for understanding endemic infections where there is pre-existing immunity that may be heterogeneous between demographic groups and across geographic space.

In conclusion, this study describes previously unreported characteristics of mobility networks in Dhaka and Bangkok, and reports important differences in the trajectories of simulated epidemics propagated over these networks. Our findings support the continued development of passively-collected mobile user data as an important tool for understanding and predicting the *a priori* risk of epidemic propagation in different cities, and for planning disease control interventions that are predicated on understanding the spatial and temporal dynamics of epidemics at the city (including, for example, the distribution of public health resources for case detection and testing).

## Data Availability

Data referred to in the manuscript are available with the supplemental information.

## 6 Acknowledgements

This work was supported by the United States National Institutes of Health (T32AI007061 to TSB and R35GM124715 to COB.)

## 7 Supplemental Methods

### 7.1 Characterizing the spatial distribution of population distribution in cities

We calculate measures of dispersion for population density distributions (Gini coefficient, entropy, and relative standard deviation) using standard approaches described in [10]. We calculate the “spatial dispersion index” (“spreading index” in [10]) as follows:

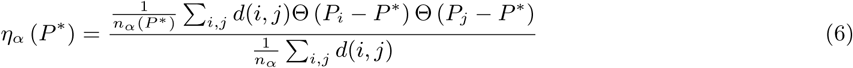

where *P* ^*^ is a threshold value above which cells in the area map are identified as having high population density. We calculated the dispersal index using two different values for *P* ^*^: (1) the mean population density and, following [cite], (2) the “Loubar” value derived from the Lorenz curve of population counts. To address possible bias introduced by the shape and size of the study area in each city, we calculated the above metrics over concentric circular areas of radius *r* centered on the map location with the highest population density in each city and report each metric across a range of *r* values

### 7.2 CDR data collection and processing

Anonymized CDR data was collected from 4415 individual network towers in the Bangkok Metropolitan Region between 1 August and 19 October 2017. We excluded two time periods from the dataset (12-13 August and 13-15 October) corresponding to two major national holidays during which total subscriber counts transiently decreased, leaving a total of 81 days in the dataset. In Dhaka Statistical Metropolitan Area, CDR data was collected from 1560 network towers. The entire data collection period in Dhaka spans 183 days from 1 April to 30 September 2017. The total number of subscribers in the Dhaka dataset increased from 22.6 million to 31.5 million during this data collection period. To minimize any potential bias introduced by this increase, and to match the duration of the data collection period of the Bangkok dataset, we used only the first 81 days of data in Dhaka dataset. Three towers in Dhaka exhibited unusual daily subscriber counts (which progressively increased during the study period) were omitted from analysis. We also identified a small number of towers (*<* 1% in both cities) that appear to have been activated during the data collection period. Specifically, these towers were found to have zero total subscribers until a discrete day, and from that point forward had large, non-zero subscriber numbers. We calculated the mean number of trips originating from these specific towers using only on the non-zero, post-activation days in each dataset.

As described in *Methods*, we consider two origin-destination matrices, 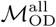 and 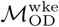 specifying connectivity between nodes in the Dhaka and Bangkok city-level mobility networks. Mantel testing, performed using the untransformed origin-destination matrices as “similarility” rather than “distance” matrices, indicated statistically significant correlation between between 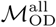 and 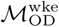 for Dhaka and Bangkok (*p <* 0.005 for all comparisons with n=1000 permutations). Community structures estimated using either 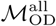 or 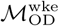 are broadly similar: similarity and dissimilarity metrics (including normalized mutual information, Rand Index, and variation of information) for community structures estimated using the two different origin-destination matrices are similar those estimated for multiple Infomap estimates using the same origin-destination matrix, i.e. the magnitude variation between community structures estimated 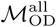 or 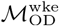 is similar to the variation inherent to all Infomap-based estimates of community structure (Figures S2 & S3). The geospatial and statistical distribution of eigenvector centrality values estimated from 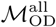 and 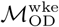 are broadly similar (Figures S4 & S5).

Lastly, we evaluate whether the observed differences in estimated community structures between Dhaka and Bangkok could be attributable to unmeasured connectivity between nodes within city-level commuting networks and unobserved nodes outside each data catchment area (i.e. the BMR and DMSA shown in Figure 2 A). Specifically, to examine whether the geographically-unconstrained communities observed in Dhaka result from connectivity with unobserved nodes outside the DMSA, we test whether the geographic contiguity of the Bangkok community structure is lost if increasing numbers of nodes are excluded from the origin-destination matrix, effectively creating an increasing number of unobserved nodes. Estimated community structures in Bangkok retain strong geographic contiguity (measured as the proportion of a node’s immediately adjacent neighbors that belong to the same community and reported as the average of this value for all nodes) even when the network is restricted to nodes within 15 km of the city center (Figure S7).

## 8 Supplemental Figures

**Figure S1:**
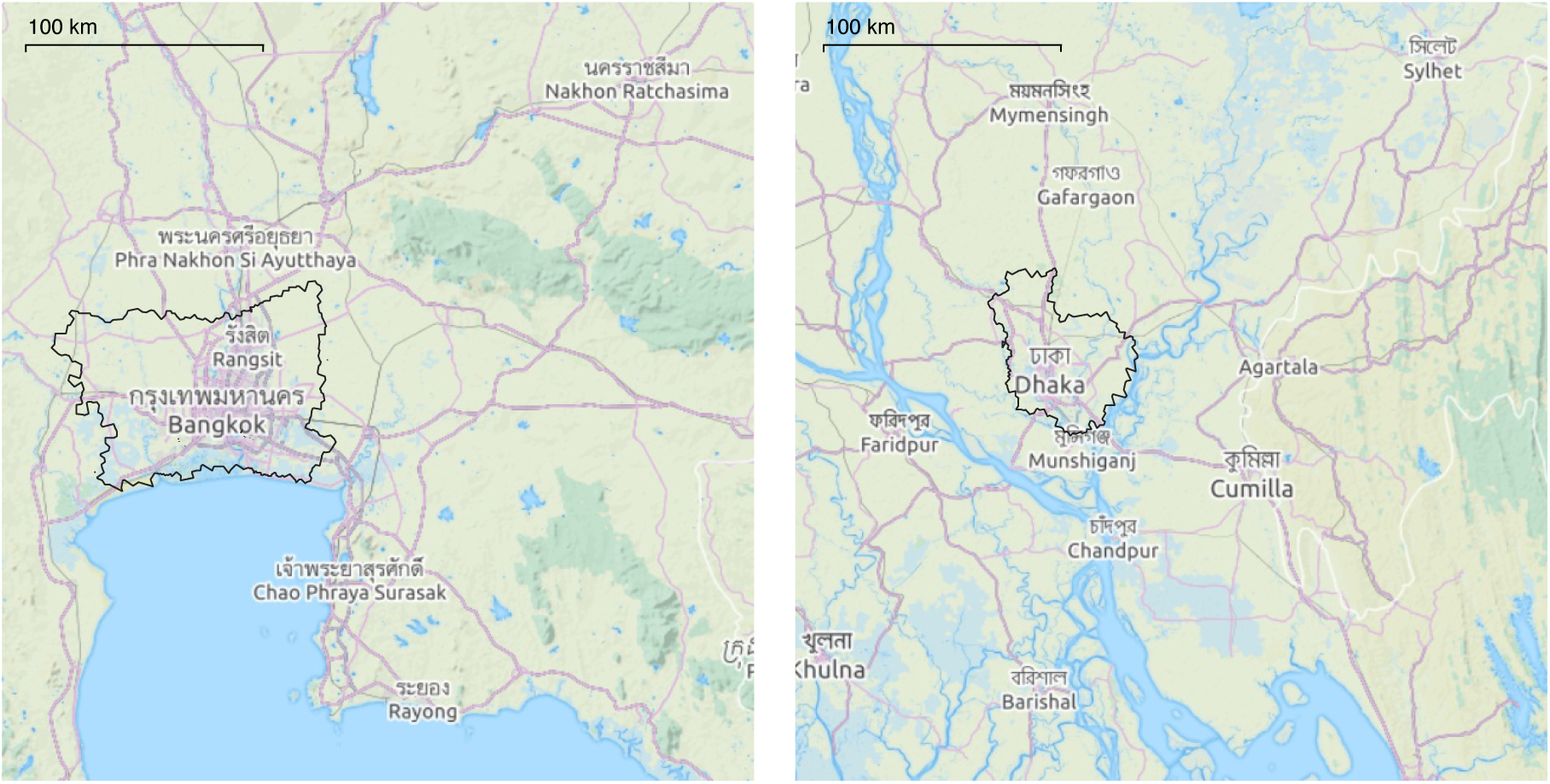
Data collection areas for the Bangkok Metropolitan Region (BMR, left) and the Dhaka Metropolitan Statistical Area (DMSA, right). Mobile phone-associated movement data was collected for mobile network within the BMR and DMSA (demarcated in black).

**Figure S2:**
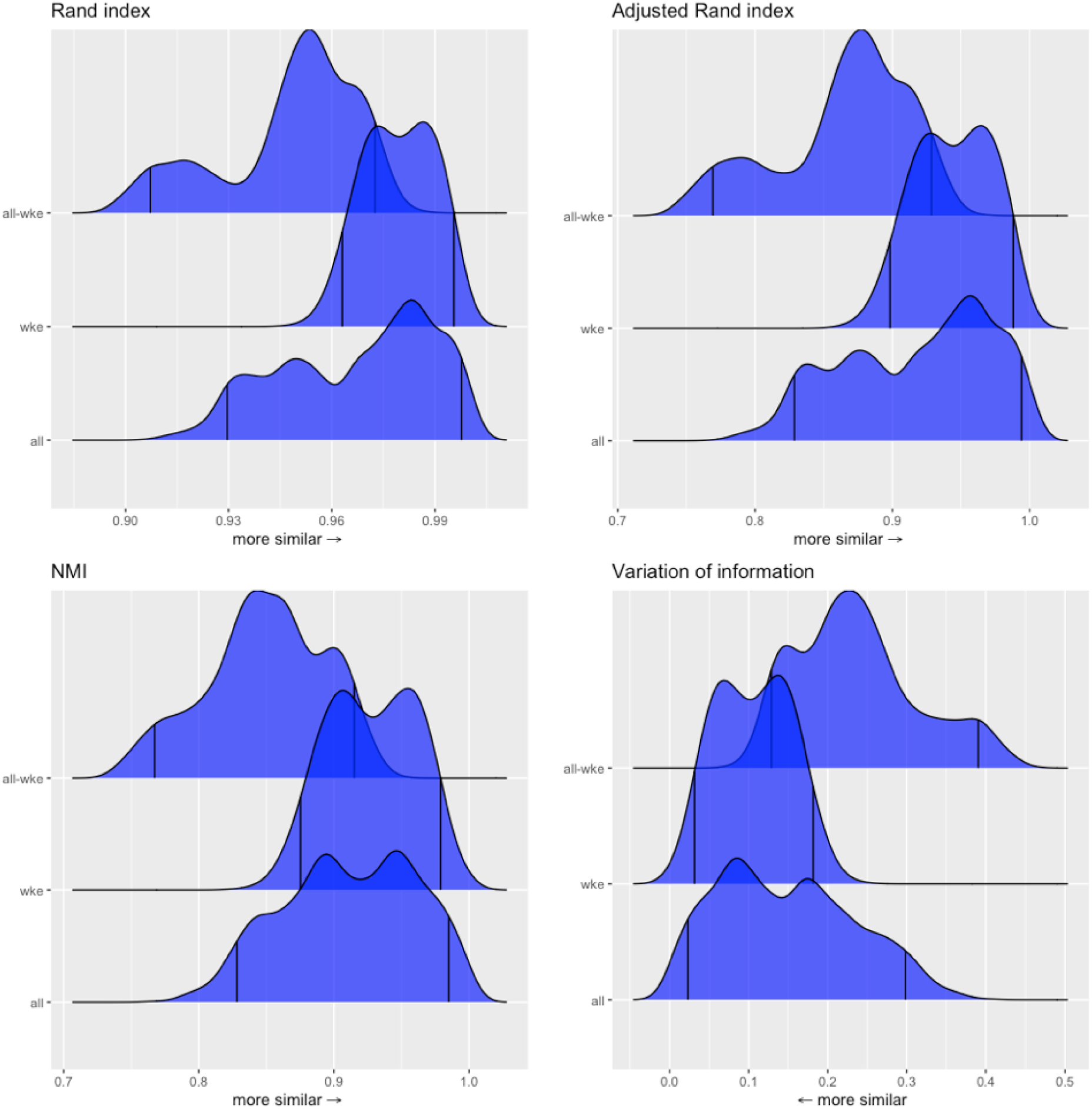
Infomap-generated community structures estimated using the same or different data sources in Dhaka. Distributions labeled “all” in each panel show the normalized mutual information (NMI), Rand index, or variation of infomation values for 1225 unique pairwise comparisons between n=50 unique Infomap-generated community structures estimated using 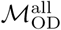. Likewise, distributions labeled “wke” show the distribution of these metrics for pairwise comparisons between estimates based on 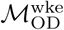. Distributions labeled “all-wke” show NMI, Rand index, and variation of information values for 2500 unique pairwise comparisons between n=50 Infomap community structure estimates using 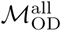 and n=50 estimates for 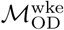.

**Figure S3:**
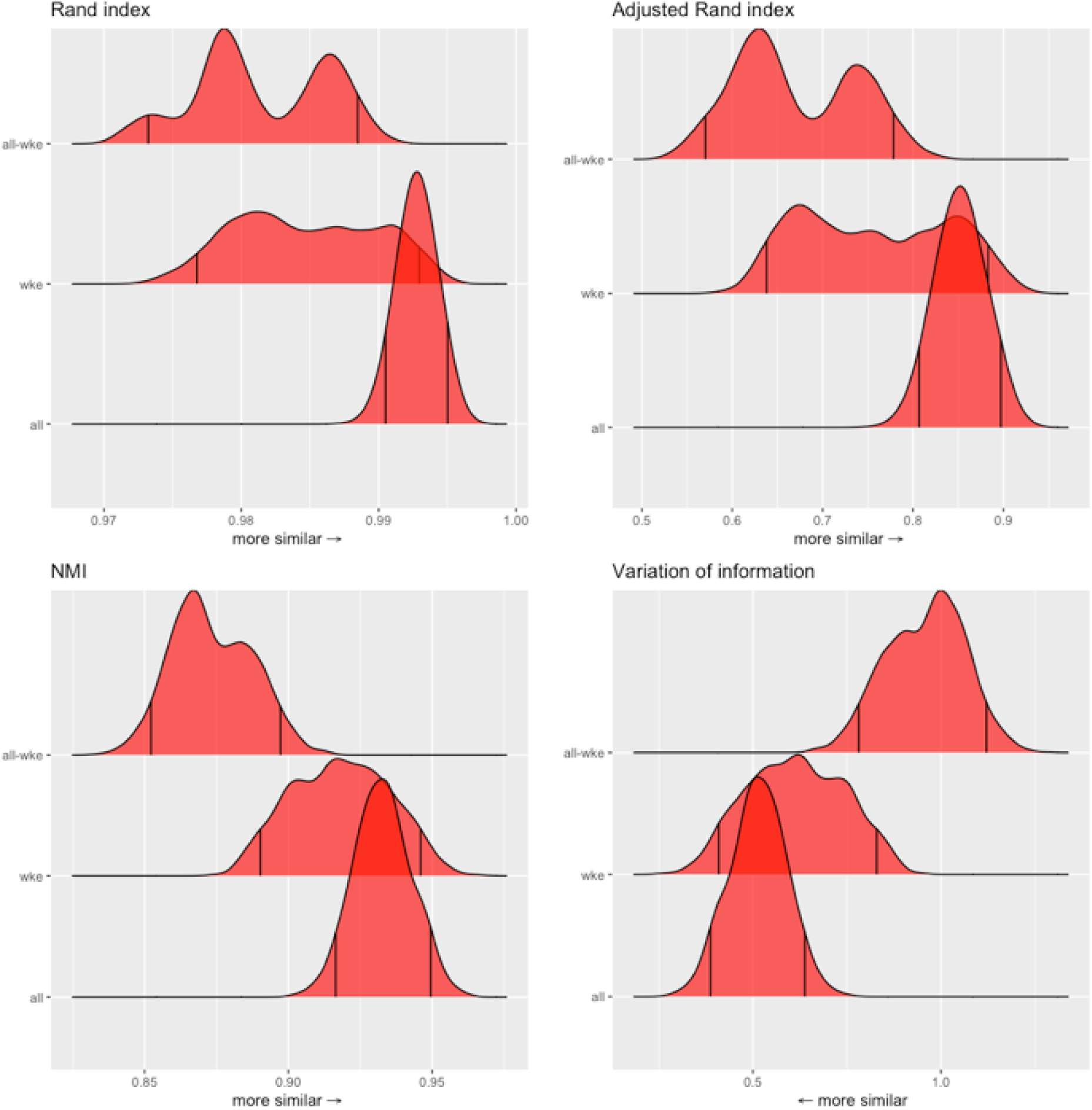
Infomap-generated community structures estimated using the same or different data sources in Dhaka. Distributions labeled “all” in each panel show the normalized mutual information (NMI), Rand index, or variation of infomation values for 1225 unique pairwise comparisons between n=50 unique Infomap-generated community structures estimated using 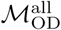. Likewise, distributions labeled “wke” show the distribution of these metrics for pairwise comparisons between estimates based on 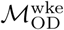. Distributions labeled “all-wke” show NMI, Rand index, and variation of information values for 2500 unique pairwise comparisons between n=50 Infomap community structure estimates using 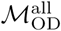 and n=50 estimates for 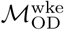.

**Figure S4:**
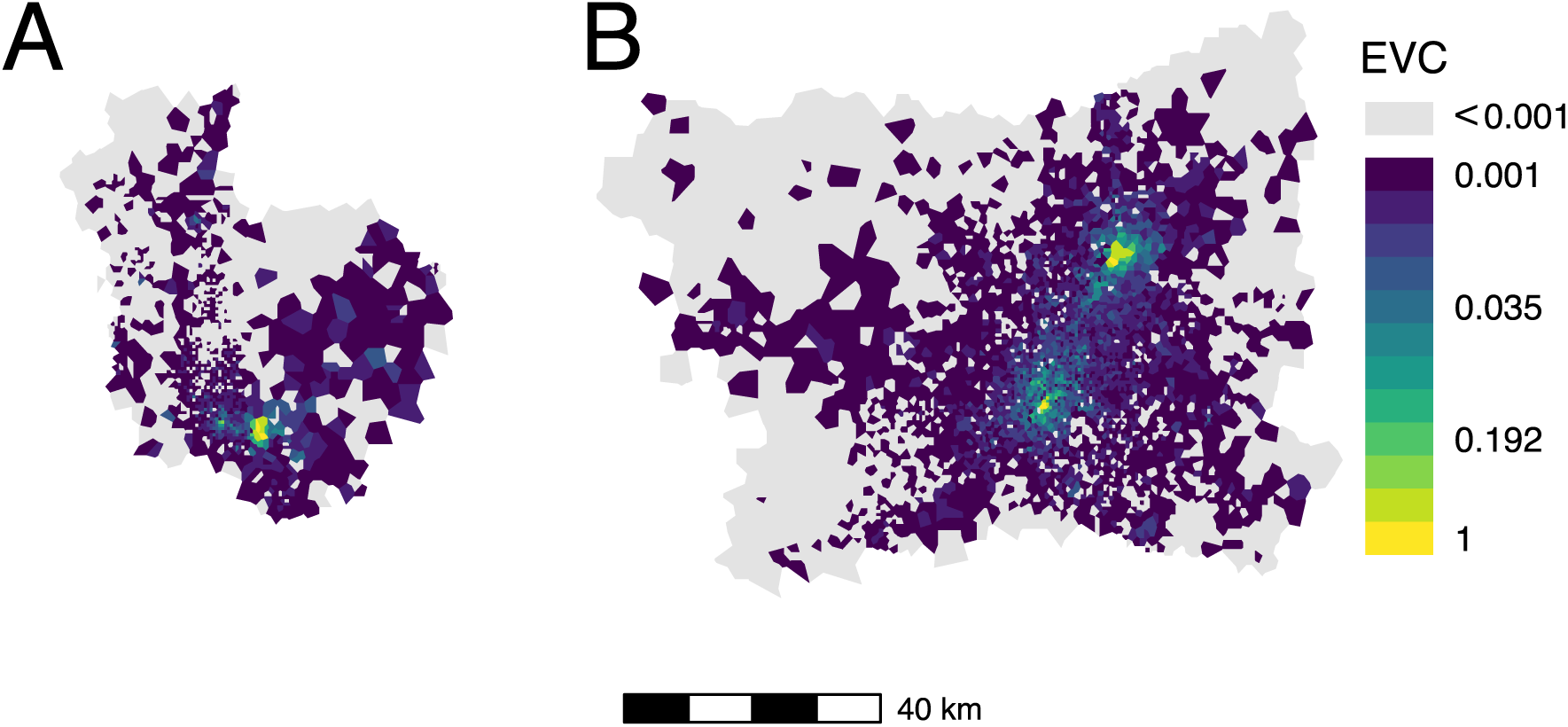
Eigenvector centrality by node for (A) Dhaka and (B) Bangkok using 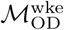 to specify the commuter network.

**Figure S5:**
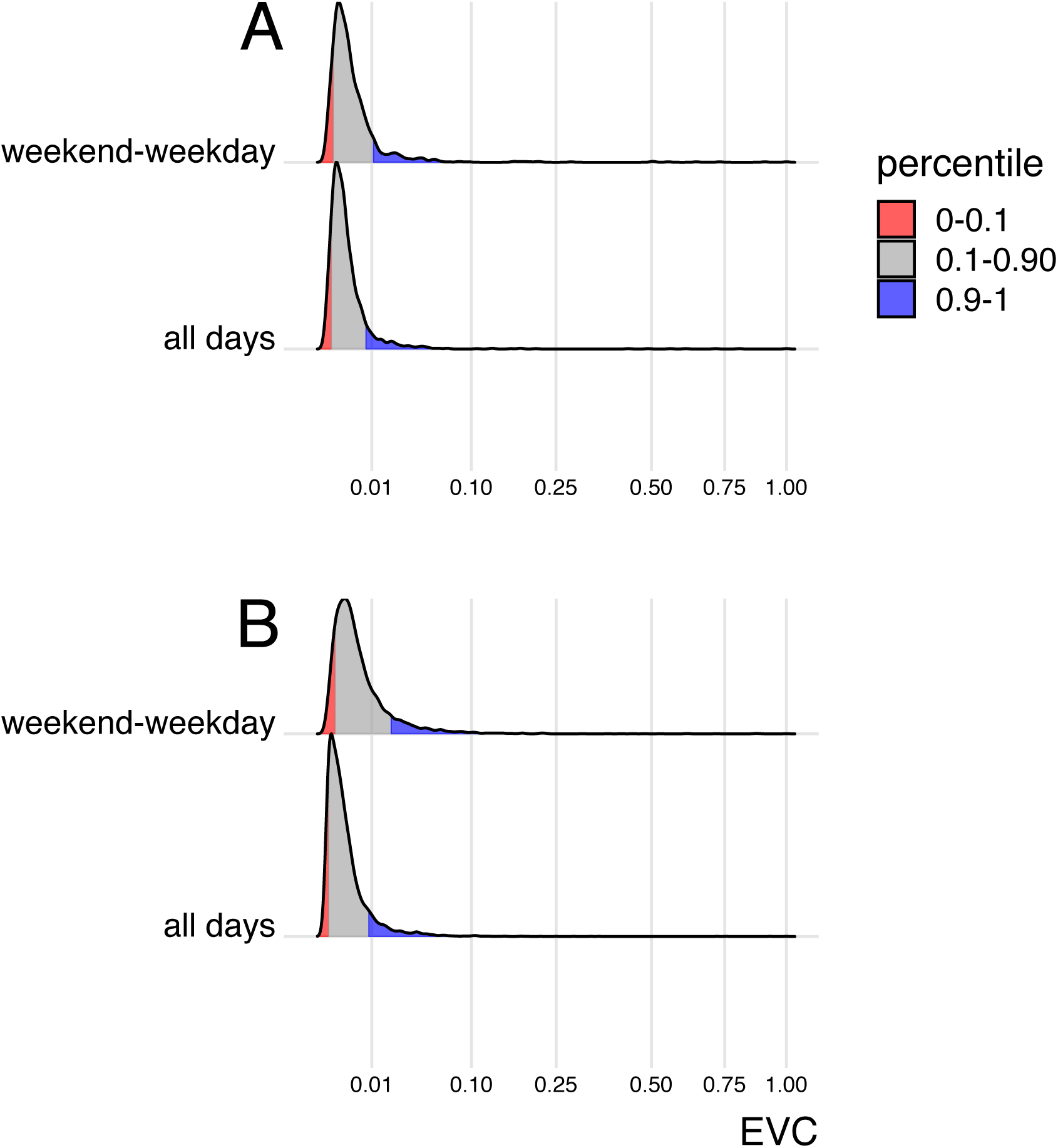
Distribution of eigenvector centrality values for Dhaka (A) and Bangkok (B), for commuter mobility networks specified by 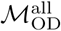 (“all days”) and 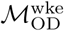 (“weekend-weekday”)

**Figure S6:**
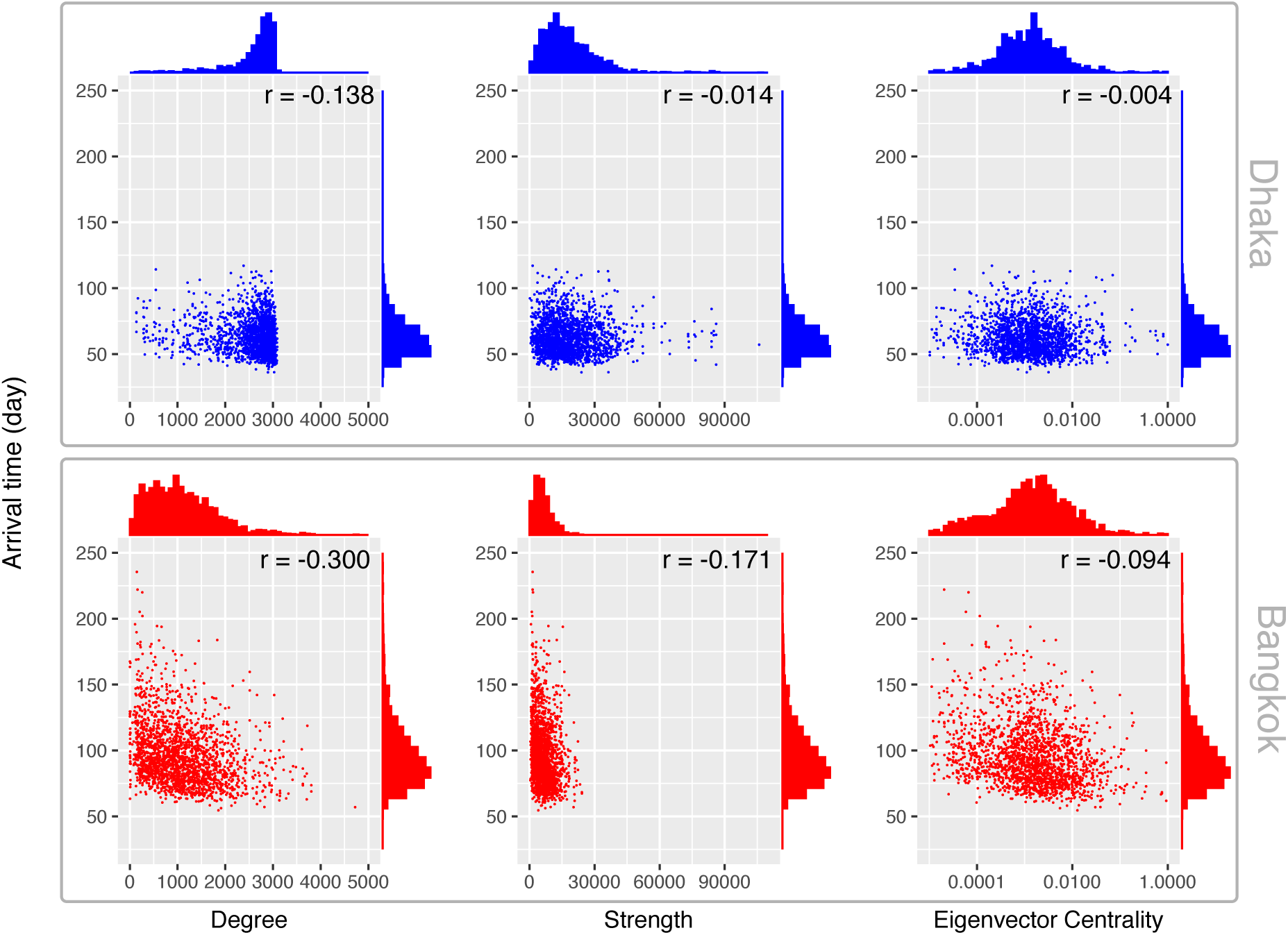
Origin node properties versus mean epidemic arrival time for Dhaka (blue, top panel) and Bangkok (red, bottom panel). Arrival time is calculated as the mean epidemic arrival time across all nodes for a given simulated epidemic. *r*: Pearson’s correlation coefficient.

**Figure S7:**
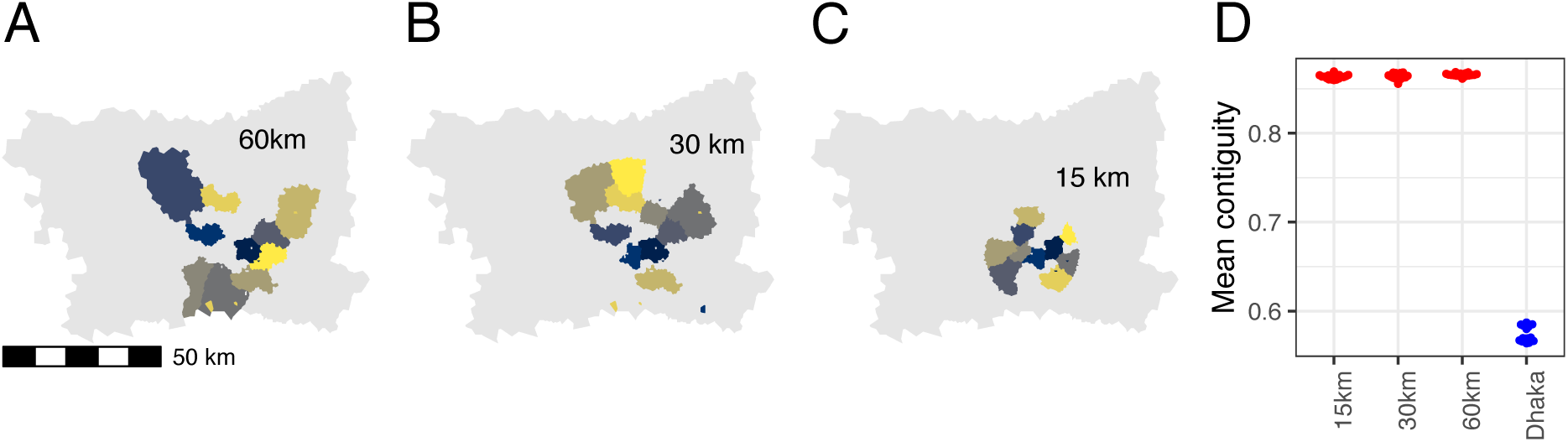
Example Infomap results, showing ten largest communities (by number of nodes), for Bangkok commuting network restricted to nodes within 60 km (A), 30 km (B), and 15 km (C) of the city center. Mean node-wise community contiguity (measured as the proportion of a node’s nearest neighboring nodes that belong to the same community) is shown for 20 independent Infomap solutions each for nodes within 60 km (A), 30 km (B), and 15 km (C) of the city center, compared to the node-wise mean contiguity for the entire Dhaka network.

**Figure S8:**
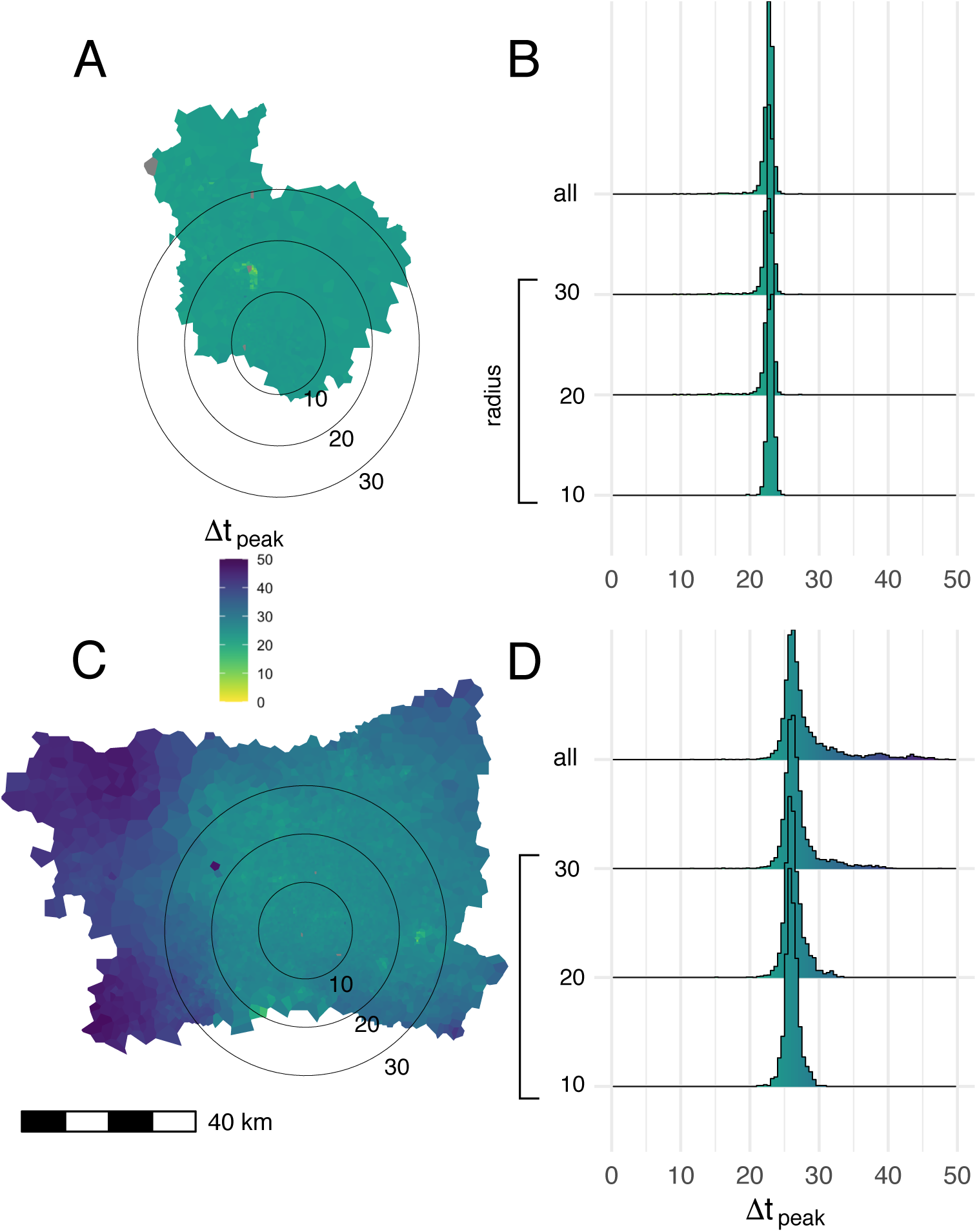
Mean time to epidemic peak for each node in the commuting network. Map polygons are colored according to the time lag in days between the time to epidemic peak in a given node and the earliest peaking node on the map (A: Dhaka, C: Bangkok). Results from 1,000 independent SEIR simulations, using parameters drawn via Latin hypercube sampling (as described *Methods*), are shown. The seed node for each simulation is drawn from a uniform distribution, where the probability that a given node is chosen as the seed is equal across all nodes. Histograms show the distribution of epidemic peak times over different geographic areas in Dhaka (B) and Bangkok (D), including nodes within concentric circles centered on most densely populated node in each city (radii: 10, 20, and 30 km) and the entirety of both study areas (“all” in panels B and D)

**Figure S9:**
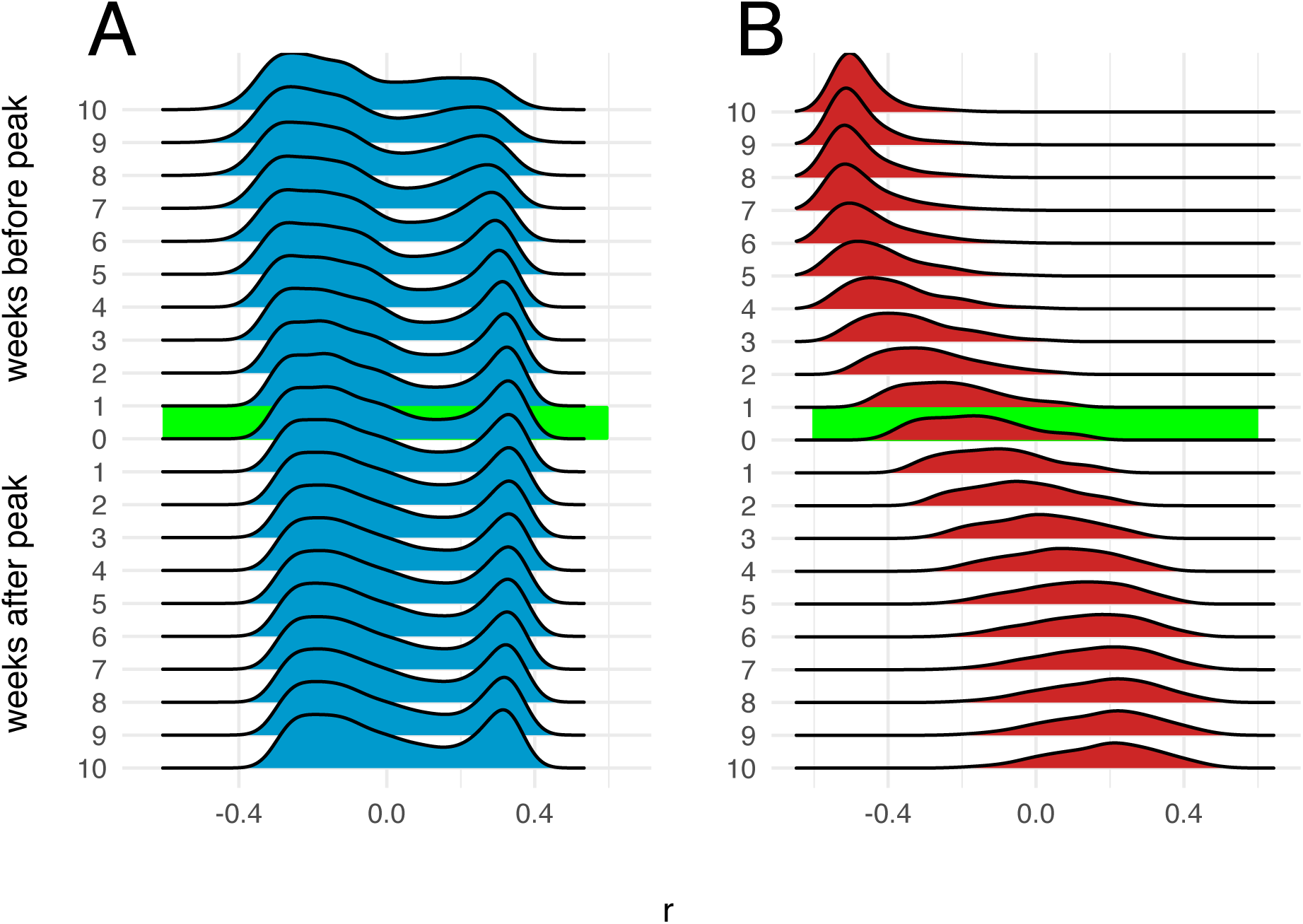
Correlation between number of infections and distance from epidemic origin node over time. Distributions of Pearson’s coefficients for correlation between number of infections in each node and nodes’ respective distances in km from the origin node, obtained from 1,000 independent SEIR simulations, are shown at different time points during simulated epidemics. The origin node for each simulation is chosen from a uniform distribution. A: Dhaka, B: Bangkok. Distribution for Pearson’s correlation coefficient at the epidemic peak are highlighted.

**Figure S10:**
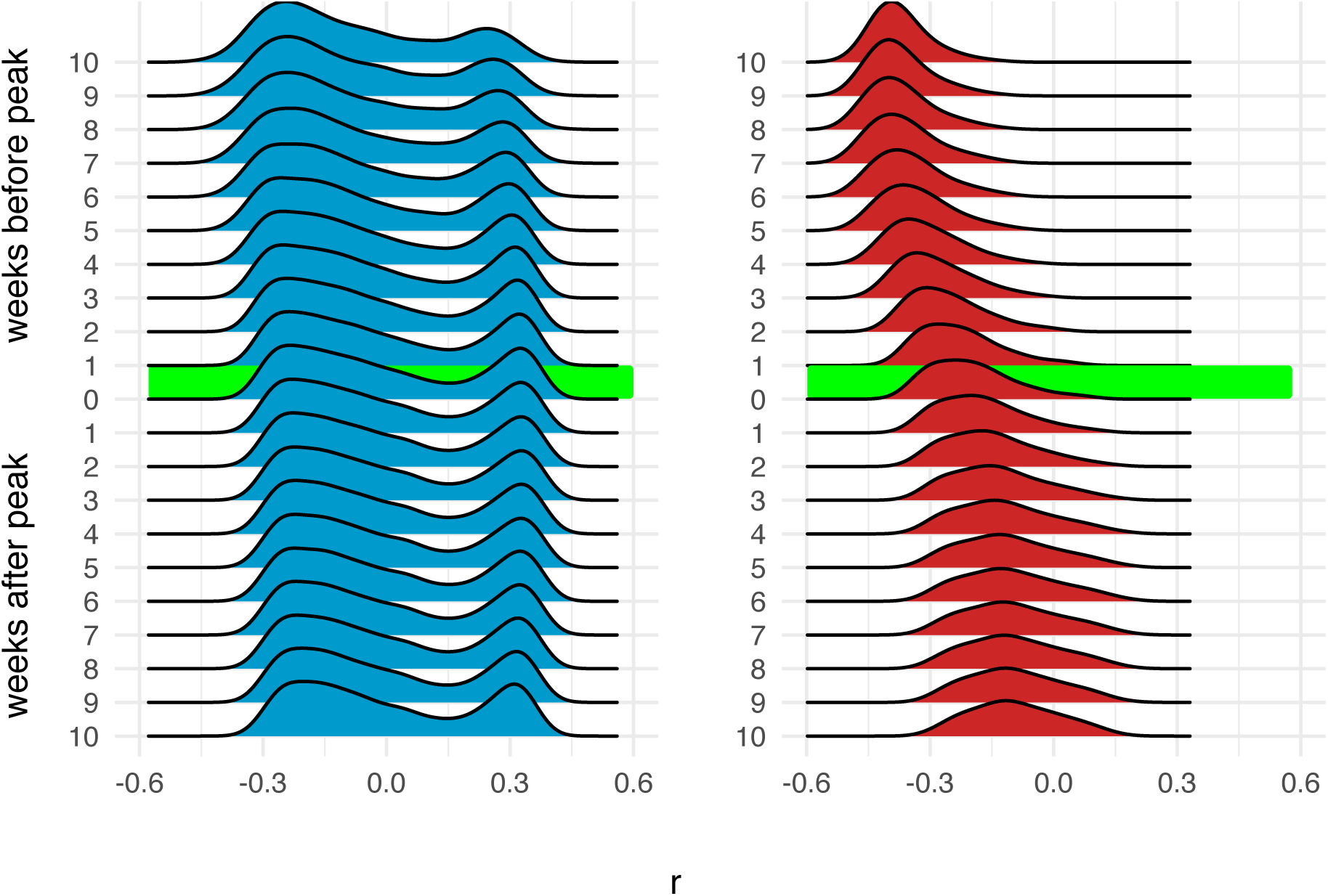
Correlation between number of infections and distance from epidemic origin node over time, for simulated epidemics propogated on 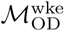. Distributions of Pearson’s coefficients for correlation between number of infections in each node and nodes’ respective distances in km from the origin node, obtained from 1,000 independent SEIR simulations, are shown at different time points during simulated epidemics. The origin node for each simulation is chosen from a multinomial distribution weighted by the total population in the catchment area (Voronoi polygon) around each node. A: Dhaka, B: Bangkok.

**Figure S11:**
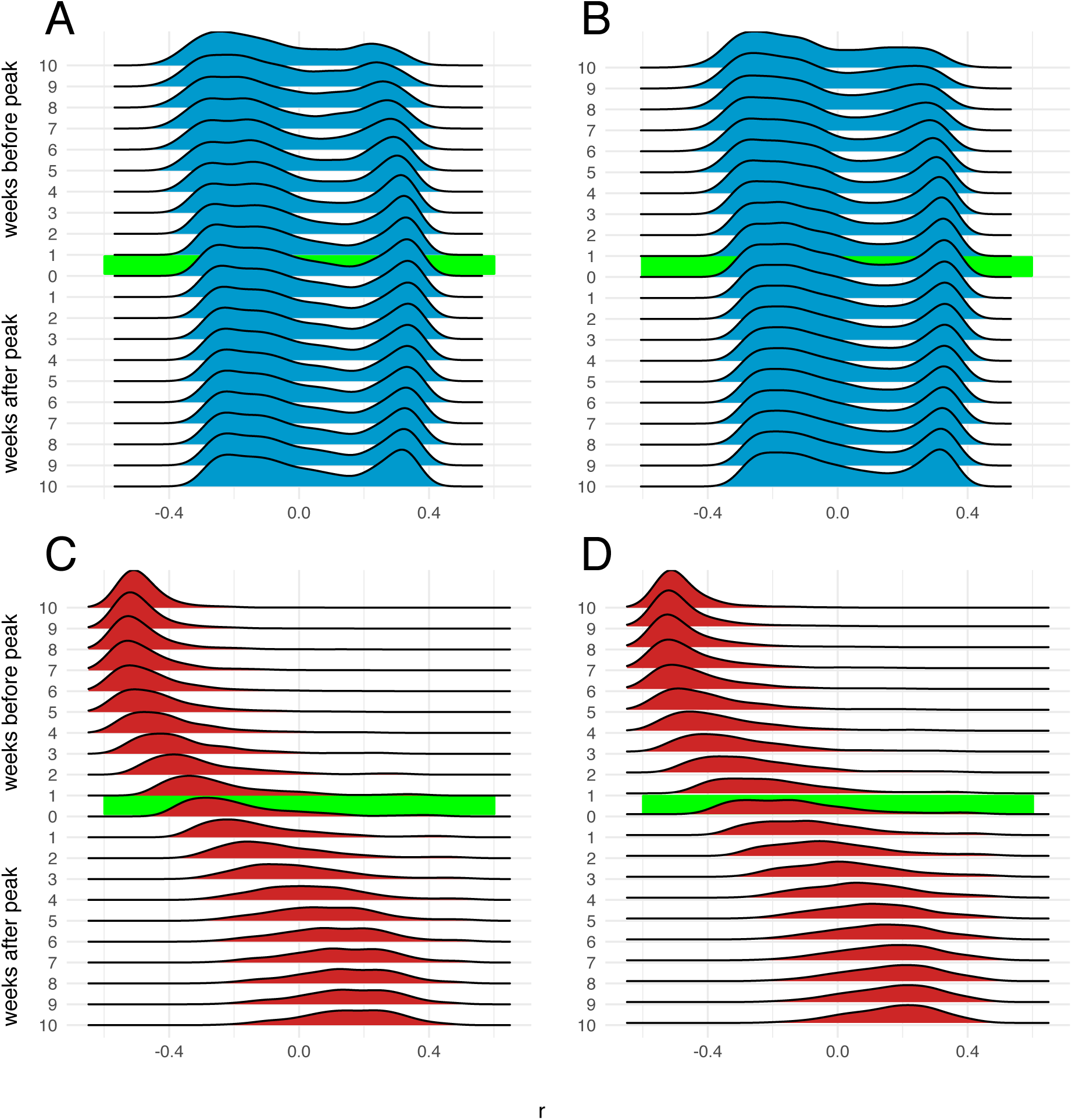
Correlation between number of infections and distance from epidemic origin node over time, restricted to nodes 60 km of the origin node. Distributions of Pearson’s coefficients for correlation between number of infections in each node and nodes’ respective distances in km from the origin node, obtained from 1,000 independent SEIR simulations, are shown at different time points during simulated epidemics. Results for Dhaka are show in panels A (origin node randomly selected from population-weighted distribution) and B (origin node randomly selected from uniform distribution), and results for Bangkok are show in panels C and D (with origin nodes selected from population-weighted and uniform distributions, respectively). Distribution for Pearson’s correlation coefficient at the epidemic peak are highlighted.

**Figure S12:**
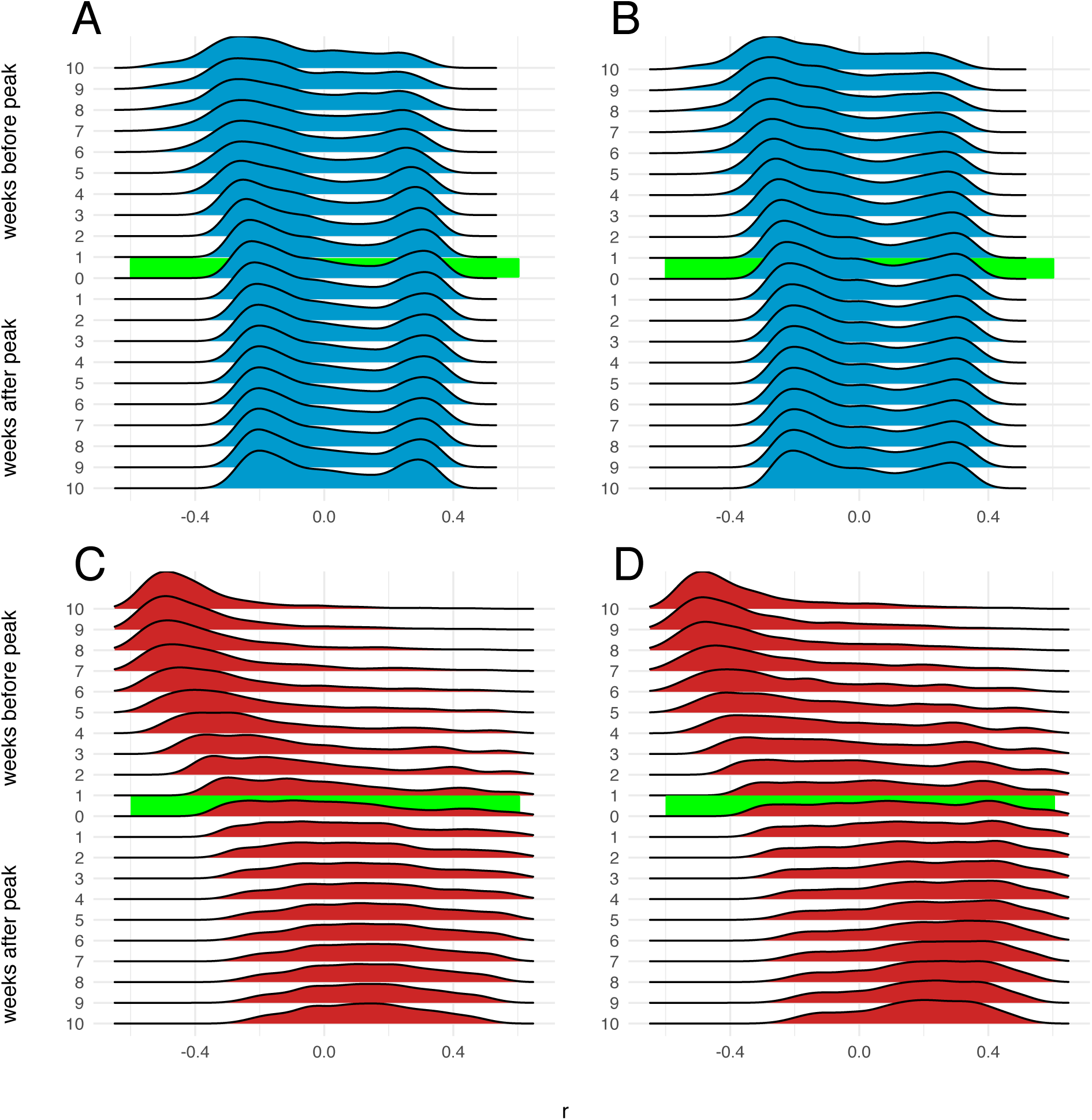
Correlation between number of infections and distance from epidemic origin node over time, restricted to nodes 30 km of the origin node. Distributions of Pearson’s coefficients for correlation between number of infections in each node and nodes’ respective distances in km from the origin node, obtained from 1,000 independent SEIR simulations, are shown at different time points during simulated epidemics. Results for Bangkok are show in panels A (origin node randomly selected from population-weighted distribution) and B (origin node randomly selected from uniform distribution), and results for Dhaka are show in panels C and D (with origin nodes selected from population-weighted and uniform distributions, respectively).

**Figure S13:**
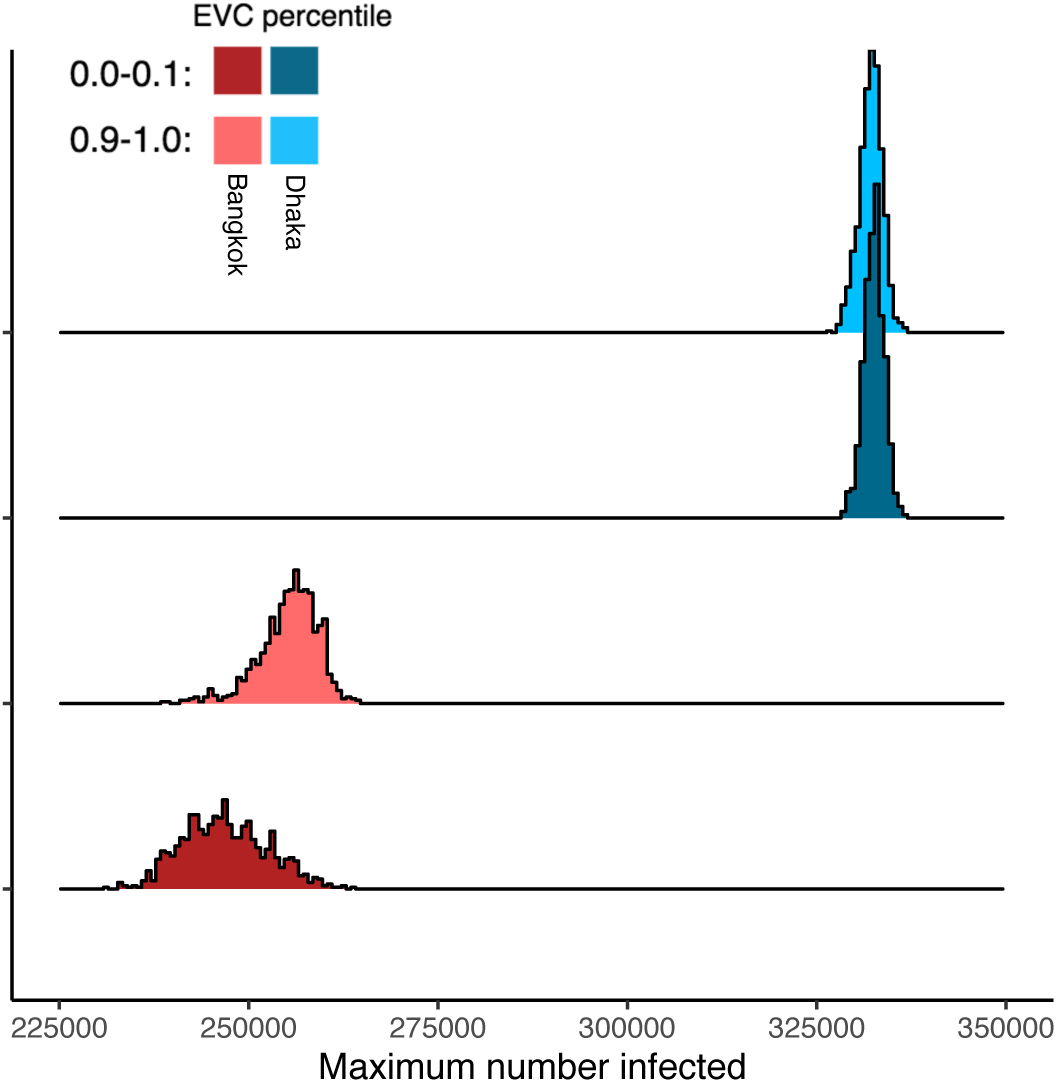
Maximum epidemic size by city and origin node eigenvector centrality. Distributions show the number of infected individuals at the epidemic peak (i.e. at *t*_peak_) for 1,000 independent simulations originated at nodes with high (tenth decile) or low (first decile) eigenvector centrality.

**Figure S14:**
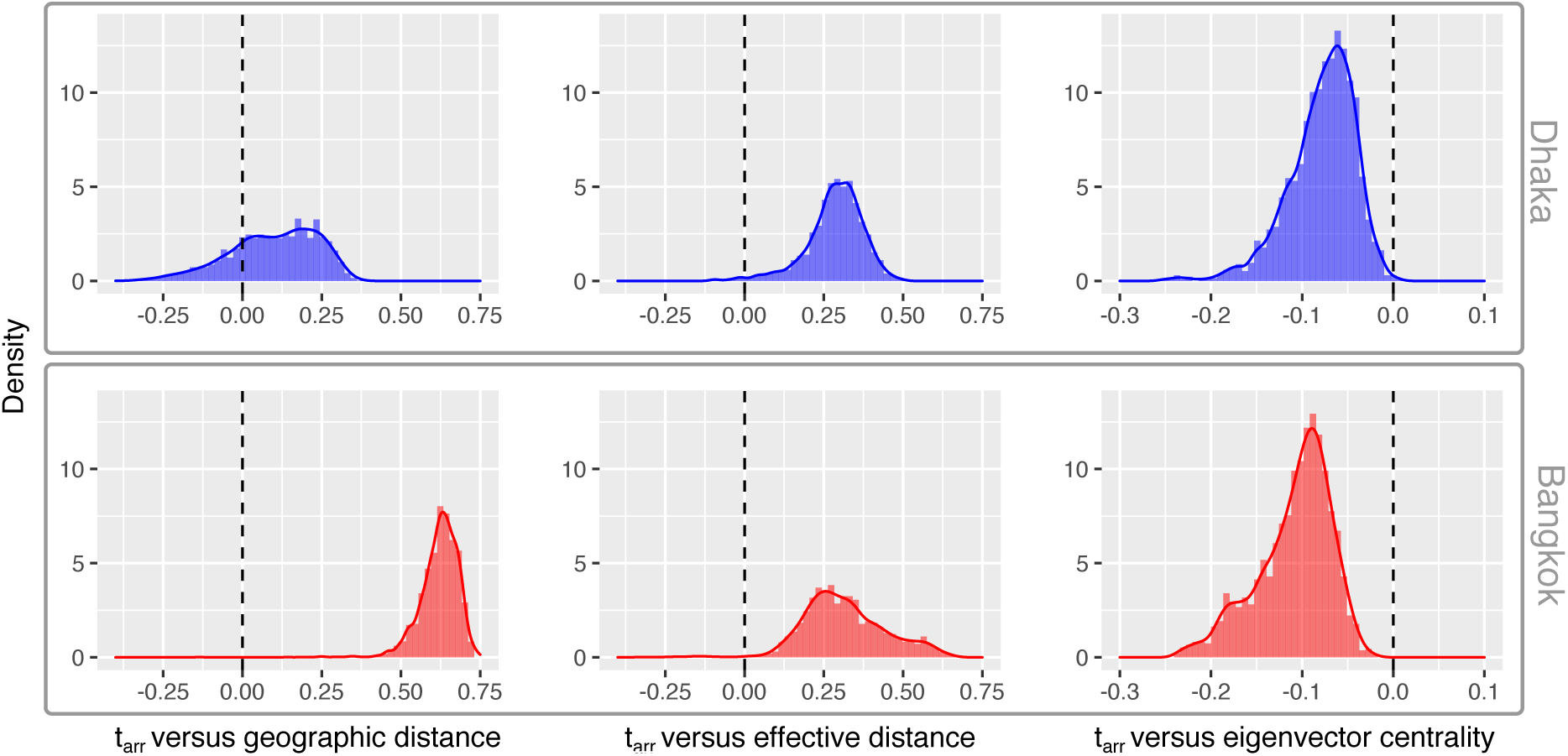
Distance from origin node and eigenvector centrality versus epidemic arrival time. Distributions of Pearson’s correlation coefficients (*r*) for each variable (geographic distance from origin node, network effective distance from the origin node ([13], and eigenvector centrality) versus the epidemic arrival time for each node are shown for Bangkok and Dhaka. These distributions are generated as follows: for each of 1000 independent simulation replicates, each seeded at a single randomly-selected node, we calculate the geographic and effective network distance between the origin node and each node in the network, and then a calculate *r* for the correlation between each of these variables and the time of the first infection in each node (arrival time, *t*_arr_) given by the simulation results. Similarly, we calculate *r* for the correlation between each non-origin node’s eigenvector centrality and its epidemic arrival time. Each distribution is generated from 1000 *r* values.

